# Sex Differences in Excess Mortality During the COVID-19 Pandemic

**DOI:** 10.1101/2025.07.15.25331003

**Authors:** Katarzyna Doniec, Jonas Scho□ley, Mine Ku□hn, Jennifer Beam Dowd

## Abstract

**Background:** The COVID-19 pandemic was a significant global mortality shock. In addition to baseline differences in mortality risk, males often face a survival disadvantage in crisis situations such as famines and epidemics. Although many countries have reported higher numbers of male COVID-19 deaths, the evolution of sex differences in mortality over the course of the pandemic, and how these differences compare using both relative (accounting for higher male baseline mortality) and absolute measures remains underexplored.

**Methods:** We examined sex differences in excess mortality from March 2020 to July 2023 using data from 33 countries in the Short-Term Mortality Fluctuations dataset. We estimate both absolute (male excess mortality – female excess mortality) and relative (male P-score – female P-score) sex differences in COVID-related mortality. We analysed the sex gap monthly and across three pandemic phases: pre-vaccine, post-vaccine, and endemic.

**Results:** In most countries, absolute male excess mortality exceeded that of females during the pre-vaccine phase of the pandemic. While the overall male disadvantage in total excess deaths gradually declined throughout the pandemic, it remained more pronounced in countries with delayed mortality peaks. The evidence for relative male disadvantage was weaker overall, and this gap further narrowed during the endemic phase. In some contexts, such as Nordic countries, women even experienced higher relative excess mortality later in the pandemic. Sex differences in absolute excess mortality tended to grow with age, but there was no clear pattern in the relative sex differences across age groups.

**Conclusions:** While previous research has highlighted a male disadvantage in COVID-19 mortality based on absolute death counts, our study shows that this pattern was concentrated in the pre-vaccine phase and declined over time. Relative increases in mortality were often similar between sexes and, in some cases, greater among women. These findings suggest that sex differences in COVID-19 mortality were more varied than commonly assumed and underscore the importance of using both absolute and relative measures to assess the impact of health crises on population subgroups. Finally, our results suggest that COVID-19 did not produce lasting shifts in pre-pandemic sex differences in mortality.

## Introduction

In modern populations, females generally have lower all-cause mortality rates than males. These sex differences in mortality have been narrowing in recent decades in high-income countries ^1–5^, a trend that may have been disrupted by the COVID-19 pandemic. Globally, higher male COVID mortality was observed early in the pandemic ^6,7^. Historically, males often have a survival disadvantage during crises such as famines and epidemics ^8^, often attributed to both biological and sociocultural factors.

For COVID-19, there are hypothesised sex differences in immune response and biological susceptibility ^9–17^. For example, women tend to have stronger and longer-lasting immune responses than men, partly due to hormonal factors like oestrogen and X-linked genetic traits, which may offer them an advantage in fighting infection. Additionally, social, occupational, and behavioural differences may have influenced exposure to the virus ^18,19^. Indeed, previous work has shown a male disadvantage in COVID-19 mortality and that the magnitude of sex differences varied by age ^20–25^ and region ^19,20,23,26–32^.

Previous studies on sex differences in COVID-19 mortality have several limitations. First, most previous studies focused on data from only the early stages of the pandemic. How sex differences evolved over the course of the pandemic and the potential effect on long-term sex differences in mortality^33^ is less known. Biological and behavioural susceptibility to COVID-19 likely evolved over time in ways that could impact sex differences but are hard to predict a priori. On the one hand, vaccines and improved treatments could reduce male biological disadvantage. On the other hand, differential uptake of COVID-19 prevention measures, such as mask-wearing, social distancing, and vaccines, could drive persistent or growing male disadvantage ^34,35^.

Second, previous studies estimated sex differences using absolute numbers of COVID-19 deaths or mortality rates. But since men have higher baseline mortality at each age, any proportionate increase in the risk of death would naturally result in more male deaths. For this reason, it is perhaps not surprising that males experienced a higher **number** of COVID-19 deaths. However, a higher number of deaths alone doesn’t tell us whether men are more susceptible to the SARS-CoV-2 virus itself, for example. In contrast, relative measures of sex differences ––such as whether one sex is seeing a higher *percentage* increase in deaths compared to expected levels–– can tell us whether sex differences in COVID-19 mortality are worse than for other causes of death. While there is no “correct” metric for quantifying sex differences, a comprehensive assessment of health inequalities can benefit from considering both absolute and relative perspectives ^36–40^.

Third, most previous studies of sex differences in COVID-19 mortality rely on data on official COVID-19 deaths. However, variations in testing practices and the accuracy of recording COVID-19 as a cause of death make cross-country comparisons challenging ^41^. By contrast, all-cause mortality is more reliably recorded and offers better comparability ^42–44^. Furthermore, examining excess mortality—the difference in observed versus expected all-cause mortality based on previous trends— allows us to contextualize the sex differences in COVID-19 mortality within pre-pandemic sex differences in each country.

We address these limitations by conducting the first comprehensive examination of trends in sex differences in excess mortality from March 2020 to July 2023, using data from 33 high-income countries. Specifically, we estimate both sex differences in excess mortality death rates and percent excess deaths by month and examine these trends across three key phases of the pandemic: pre-vaccine (March 2020 – April 2021), post-vaccine Delta/Omicron (May 2021 – April 2022), and endemic (May 2022 – June 2023). Additionally, we explore how these sex differences vary by age and country.

## Data and Methods

We used the Short-Term Mortality Fluctuations (STMF, 2025) dataset for 33 countries to estimate sex-specific all-cause excess monthly mortality between March 2020 through July 2023. We used excess mortality as a proxy for COVID-specific mortality. This approach has previously been used to estimate the death toll from other mortality crises ^45–47^. While excess mortality may also pick up indirect deaths from non-COVID causes during the pandemic, recent evidence from the US suggests that a large fraction of non-COVID excess deaths were likely undercounted COVID deaths ^48^. We estimated the expected (baseline) deaths for each week using the 7 years from 2013 through 2019 as the reference period from which to extrapolate a counterfactual non-pandemic trend.

Population denominators were sourced from the Human Mortality Database (2025). Person-weeks of exposure were calculated by cubic interpolation of the January 1st population over the weeks of a year, followed by integration of the cubic spline over single weeks. Age-specific person-weeks of exposure were linearly extrapolated by up to two years if no estimates were available.

We used a Generalized Additive Model (GAM) with smooth seasonality and a log-linear time trend to estimate expected deaths. The model was defined as

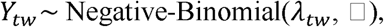

with

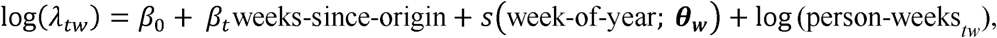

where *Y_tw_*, –– the number of deaths in week-of-year *w* at time *t*, measured in weeks since 2013 –– is distributed according to a Negative Binomial distribution with expected number of deaths λ*_tw_* and overdispersion □. Expected deaths were modelled via a log-linear long-term trend and a smooth function of week of year, *s*, implemented as cyclical cubic splines. Logged person-weeks of exposure were added as an offset to model on a death rate scale. The model was fit separately for each country, sex, and age group (0-15, 15-64, 65-74, 75-84, 85+).

Expected deaths were derived by evaluating λ*_tw_* over the period from March 2020 through July 2023. We aggregated weekly expected and observed deaths to months and across three pandemic periods: pre-vaccine (March 2020 – April 2021), post-vaccine Delta/Omicron (May 2021 – April 2022), and endemic (May 2022 – June 2023).

To quantify uncertainty in our estimates, we performed simulation-based inference based on sampled expected deaths from the Negative Binomial predictive distribution. Statistics of interest were calculated for each sample. We present median estimates along with 95% prediction intervals.

All-cause excess deaths were derived by subtracting the expected death counts from the observed death counts, taken directly from the STMF data. We left-censored excess deaths at 0 to ensure a unique interpretation of sex differences in excess deaths, e.g. a positive sex difference in excess deaths always marks higher excess deaths among males and not a smaller mortality deficit among males. Thus, we defined excess deaths, by sex, country, age, and period as the following:

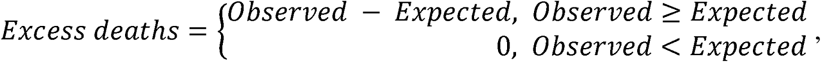

and the excess death rate (per 100,000 person-years) as

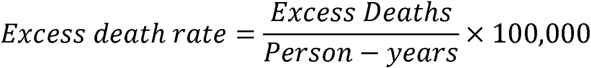

When reporting results for all age groups combined, we applied direct age standardization using the 2013 European Standard Population (Eurostat, 2013). Across age groups *x* we defined

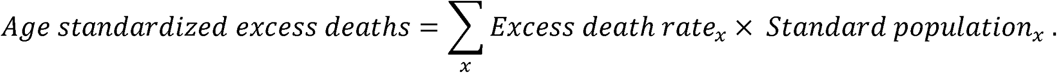

As a relative measure of excess death, we calculated the so-called P-score, the percentage difference between reported and expected deaths, as

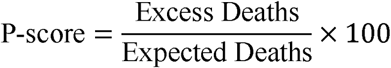

Excess mortality estimates were used to calculate both absolute and relative measures of the sex gap. The absolute gap is the difference between male and female all-cause excess death rates. The relative gap is calculated as the percentage point difference between male and female excess mortality P-scores. Further, we calculated expected (under pre-pandemic trends) and observed sex ratios in all-cause mortality.

Due to the large number of country-age group combinations, we discuss age-specific results for six countries reflecting a range of pre-pandemic sex differences in mortality, sociocultural contexts, and pandemic experiences: Bulgaria, England & Wales, Germany, Italy, Norway, and the United States. Results for the full set of countries are available in the Supplementary Information. Among high-income nations, the United States and England & Wales recorded among the highest per capita COVID-19 death rates. Italy faced high COVID-19 mortality early in the pandemic, implemented high levels of pandemic restrictions and has one of the world’s oldest populations. Norway, a high-income economy known for its high life expectancy, despite low levels of restrictions, experienced low overall COVID-19 mortality and a later onset of excess mortality. Germany, another wealthy European nation, implemented strict restrictions and experienced low excess mortality in 2020, followed by high excess in 2021-2023. Bulgaria, an Eastern European nation with large pre-pandemic sex disparities in mortality and one of the highest proportions of older adults in the world ^49^, experienced one of the highest COVID-19 mortality rates in the world.

## Results

Figure 1 shows male-to-female ratios 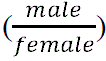 in expected (orange dots) and observed (green dots) all-cause mortality by period and country, ordered from lowest to highest expected ratios based on pre-pandemic trends. A ratio of 1 means no sex difference in the mortality rate. Ratios above 1 indicate higher male mortality; ratios below 1 indicate higher female mortality. This figure shows the range of cross-country mortality sex differences prior to the pandemic, ranging from small differences in Iceland, Norway, Sweden to large differences in the Baltic countries (Estonia, Lithuania, Latvia). In the pre-vaccine period, the male-to-female ratio of observed mortality was typically larger than the ratio for expected mortality, indicating the male disadvantage was worse than expected based on pre-pandemic mortality. This can be particularly observed for the Netherlands, Switzerland, Czechia, and Bulgaria.

**Figure 1.**
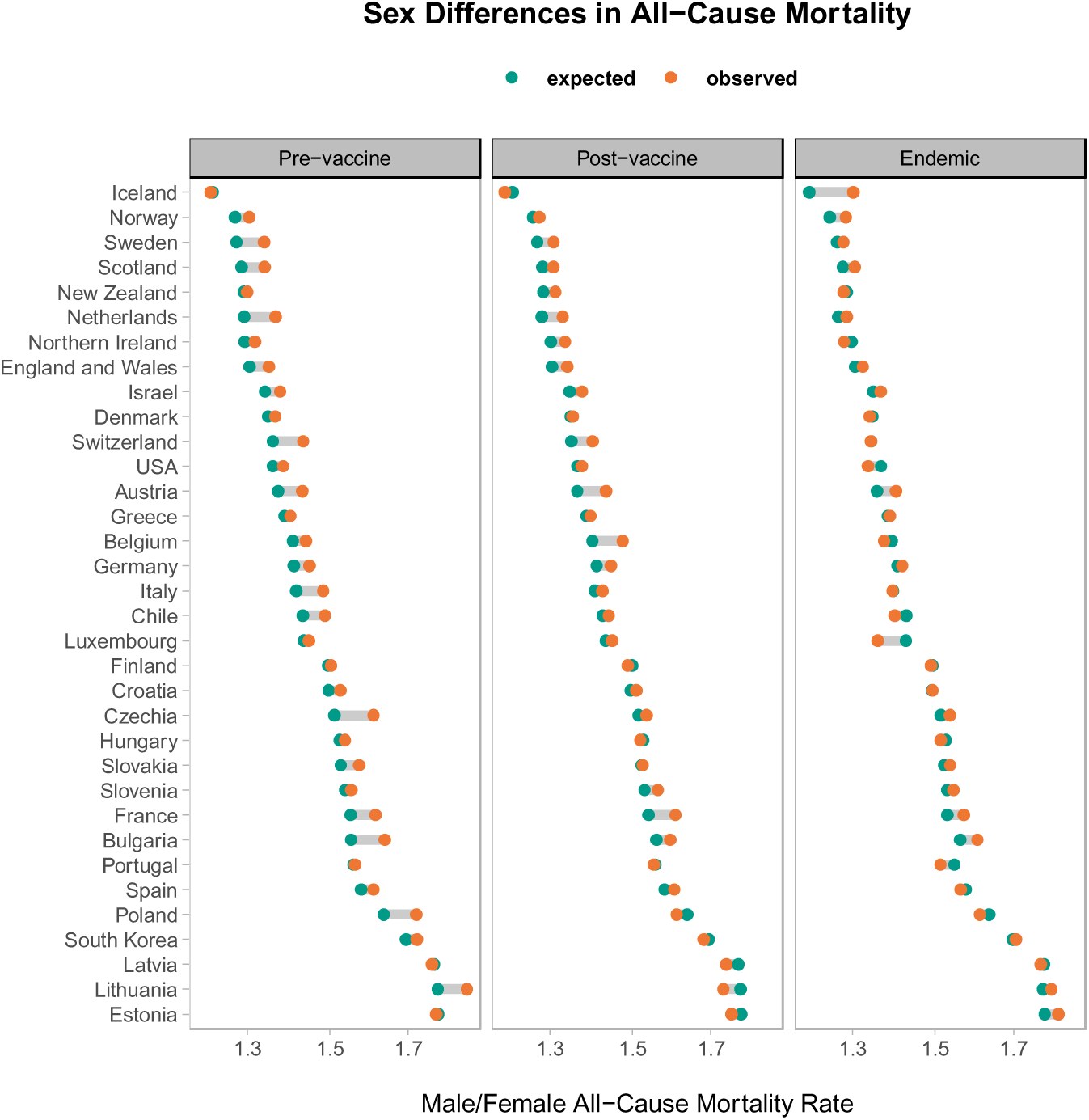
Sex ratios in all-cause mortality by country and period. **Note:** Ratio of 1 indicates no sex gap in mortality. Ratio> 1 indicates male disadvantage. Ratio<1 indicates female disadvantage.

This gap tended to decrease as the pandemic progressed, but the male disadvantage compared to expected mortality ratios persisted through 2023. In 26 out of 33 countries, sex ratios in observed mortality got smaller during the endemic phase, compared to the pre-vaccine period. Belgium, Estonia, Finland, France, and Iceland showed little or no change between the two periods. In contrast, New Zealand and Slovenia experienced an increase in sex ratios during the endemic phase, suggesting a growing male disadvantage.

Figure 2 shows monthly trends in all-cause excess mortality death rate per 100,000 person-years by sex between March 2020 and July 2023 across six countries and four age groups. Figures for the remaining countries are available in the Supplementary Information (Figures S1-S5). In general, the difference between male and female excess mortality death rate fluctuates over time depending on the overall mortality levels. In months with higher mortality, the sex difference grows, with males seeing higher absolute excess than females. During months with lower mortality, the 95% prediction intervals for male and female excess mortality largely overlap (i.e., no evidence for male disadvantage in these periods).

**Figure 2.**
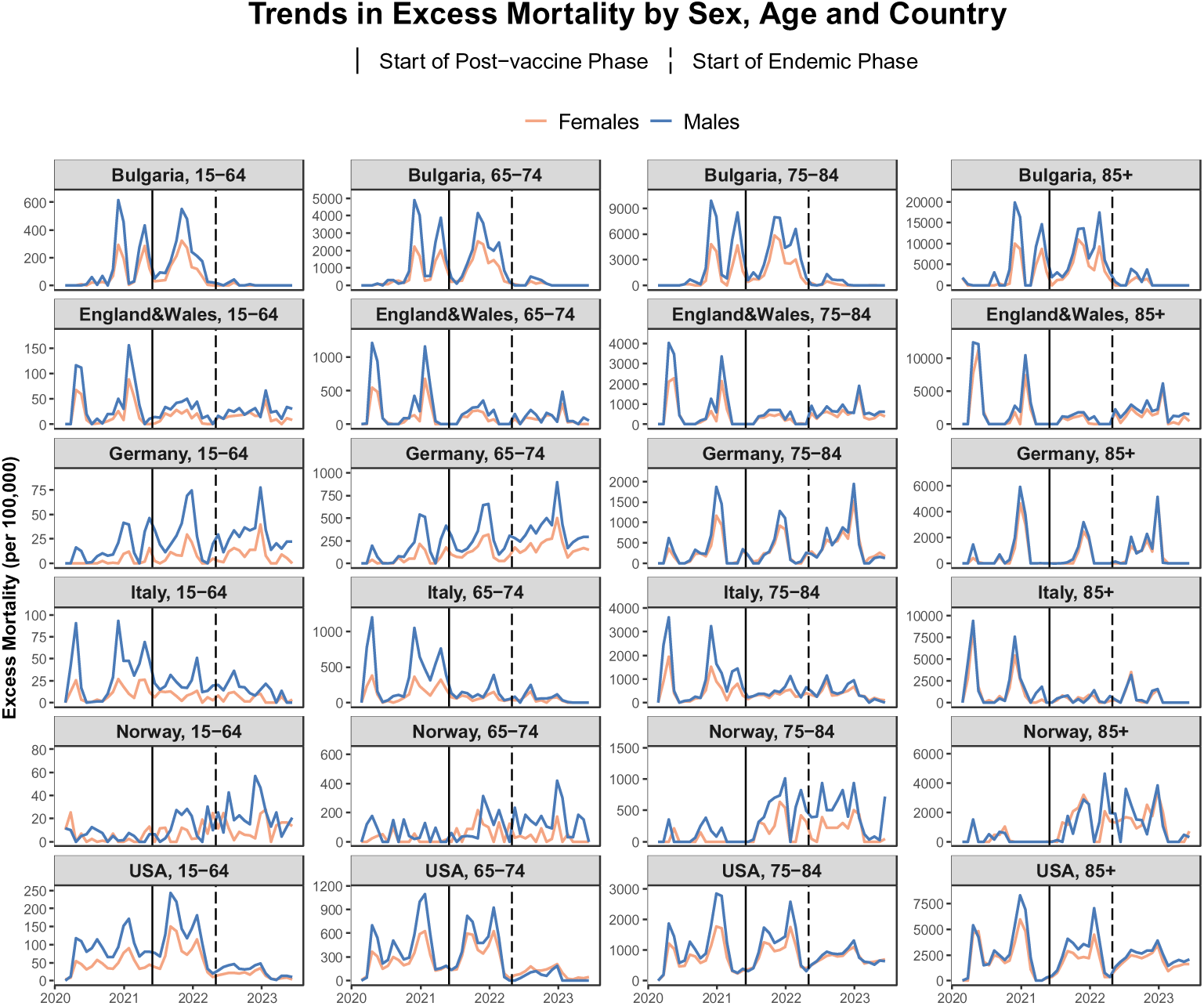
Monthly trends in excess mortality death rate (per 100,000 person-years) by sex, age, and country. **Note:** the scale of the y-axis differs for each age group and country

Within specific countries, Bulgaria and the USA experienced excess mortality peaks with a sizeable male disadvantage in both pre– and post-vaccine phases, but sex differences narrowed with lower mortality levels in the endemic phase. England & Wales and Italy saw large peaks in excess mortality in the pre-vaccine period, with a clear male disadvantage in months where mortality peaked. Germany saw mortality peaks in all three periods, but significant sex gaps were only seen under age 75. Norway saw lower mortality levels early in the pandemic, with peaks occurring later in the endemic phase. In Norway, the sex gap remained negligible in most months (i.e. prediction intervals for males and females were overlapping), including peak months in post-vaccine and endemic phases.

In Denmark, Finland, Estonia, Iceland, New Zealand and Northern Ireland, the 95% prediction intervals for male and female excess mortality overlap across all three periods (i.e., no evidence for male disadvantage).

Figure 3 illustrates monthly trends in P-scores by sex and age between March 2020 and July 2023 for the selected six countries. Results for the remaining countries are available in the Supplementary Information (Figures S6-S10). Sex-specific trends are shown as P-scores on the same percentage scale (from 0% to +80%) in all countries except Bulgaria (from 0% to +120%), which experienced higher excess mortality than the remaining five countries.

**Figure 3.**
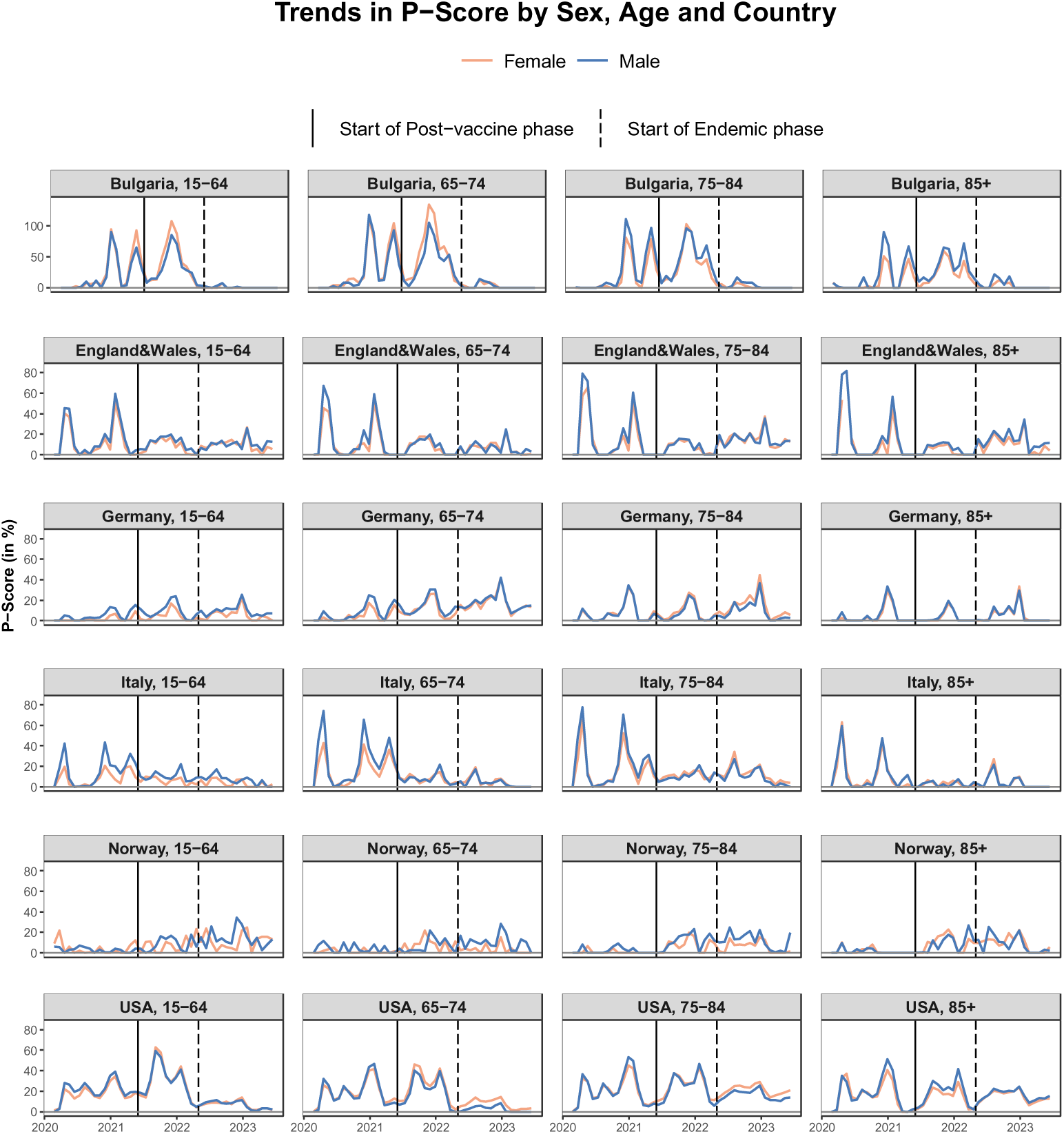
Monthly trends in excess death P-scores by sex, age and country. **Note:** the scale of the y-axis is fixed for all countries and age groups except Bulgaria Given changing biological and social dynamics during the pandemic (e.g. evolution of the COVID-19 virus, introduction of vaccines, changing levels of government lockdowns and restrictions), we next compared changes in absolute and relative sex differences in excess mortality across three distinct pandemic phases. Figure 4 shows male and female excess mortality death rates, while Figure 5 shows male and female P-scores, both with corresponding 95% prediction intervals, calculated separately for each age group and period across six countries. Additional figures for all countries, together with 95% prediction intervals for the difference between male and female estimates, are available in the Supplementary Information (Figures S11-S16 and S17-S22).

In contrast to the trends in absolute sex differences in excess mortality (Figure 2), male disadvantage in P-scores was rare, regardless of mortality levels (Figure 3), and in some cases, females had higher relative mortality increases than males (e.g., 65-74 and 75-84-year-olds in the US during the endemic period). Higher female P-scores mean that deaths for women increased by a higher percentage from the baseline (expected mortality) levels compared to those for men.

To summarize, males suffered higher absolute levels of excess mortality during pandemic peaks in most countries, especially in the pre-vaccine phase. However, trends in relative mortality by sex suggest that this gap was largely driven by higher baseline mortality among males, while percentage increases in mortality due to COVID-19 were similar across sexes for most age group-country combinations (Figure 3).

A significant male disadvantage was seen in more than half of the age group-country combinations (about 60%) during the pre-vaccine period for excess mortality death rates (Figure 4 and S11-S16 in the SI). In the post-vaccine period, this absolute male disadvantage became less common, staying significant in about half of the age group-country combinations. By the endemic period, the ‘male disadvantage either diminished, remaining significant in about one-fifth of age group-country combinations –– or disappeared altogether. There were two exceptions to this pattern: (1) countries with a delayed onset of the pandemic, such as Germany and South Korea, where the male disadvantage emerged during the post-vaccine and endemic phases for some age groups (e.g. 15-64 and 65-74-years-olds in Germany), and (2) countries with consistently low levels of male and female excess mortality death rates, such as Iceland, Finland, Luxemburg and New Zealand, where no evidence of male disadvantage was found across most age groups and periods.

**Figure 4.**
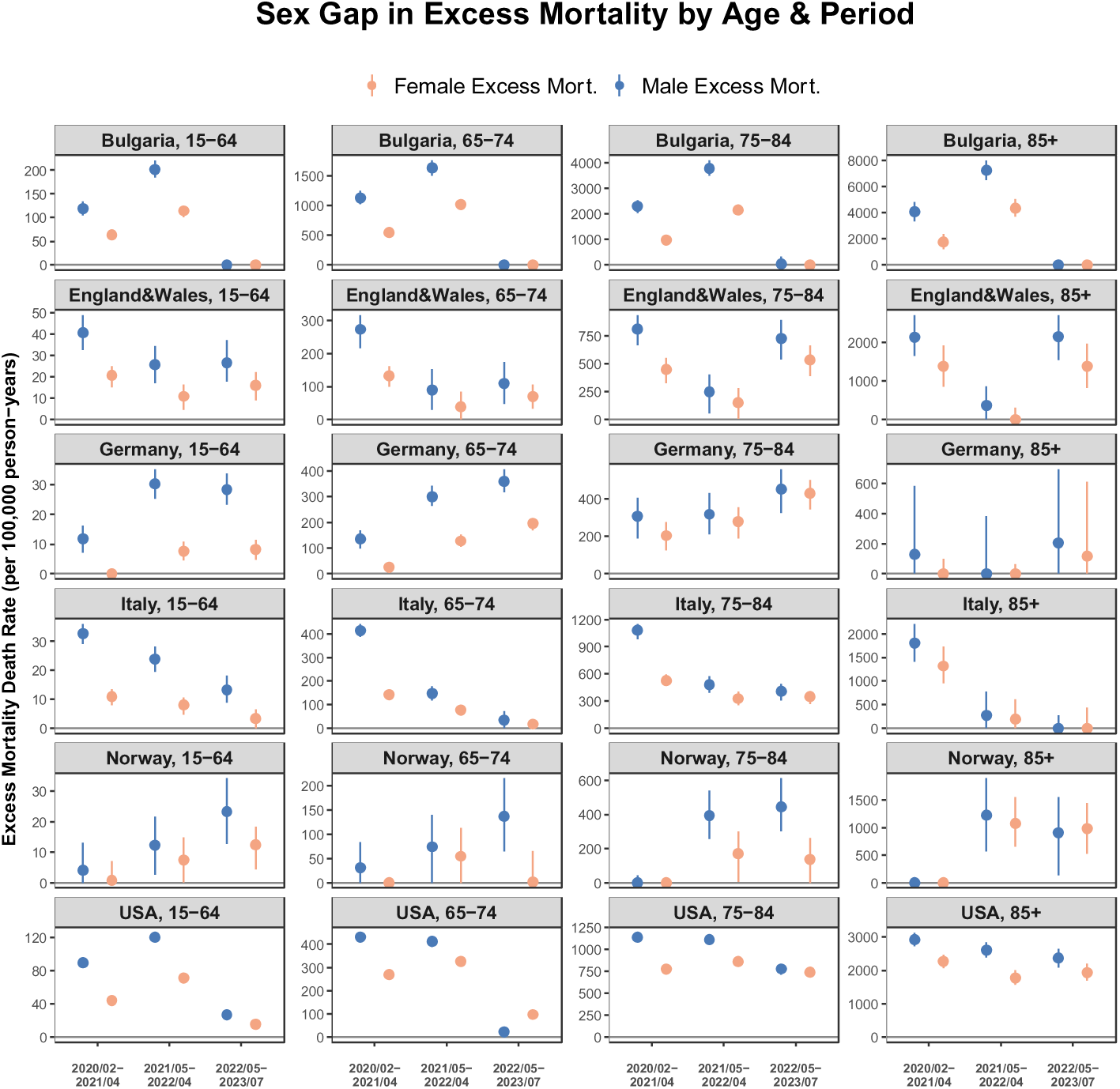
All-cause excess mortality death rate by sex, country, age group and period. **Note:** the scale of the y-axis differs for each age group and country

Significant male disadvantage during the pre-vaccine phase was much less common when looking at relative sex differences (Figure 5 and S17-S22), compared to absolute sex differences (Figure 4). For sex differences in the P-Score, in the pre-vaccine phase, a significant male disadvantage was seen in about one-quarter (28%) of age group-country combinations. In the post-vaccine phase, the relative male disadvantage further decreased, remaining significant in only one in ten (11%) of age group-country combinations. In some cases, the P-score was higher for females. For example, a female relative disadvantage emerged in the post-vaccine phase for two younger age groups in Bulgaria, for 65–74-year-olds in the USA, and 15-64-year-olds in Hungary, Lithuania and South Korea. Overall, there was little evidence of remaining relative sex gaps during the endemic phase, with a few exceptions. These exceptions included (1) countries with a delayed pandemic onset, where male relative disadvantage either continued or re-emerged in the endemic phase (e.g. 15-64 group in Germany, 65-74 and 75-84-year-olds in Norway); and (2) some cases of emerging female relative disadvantage (i.e. higher female P-score) in specific country-age combinations (e.g. 65-74-year-olds in the US, Hungary and Spain, 75-84 in Germany and the US).

**Figure 5.**
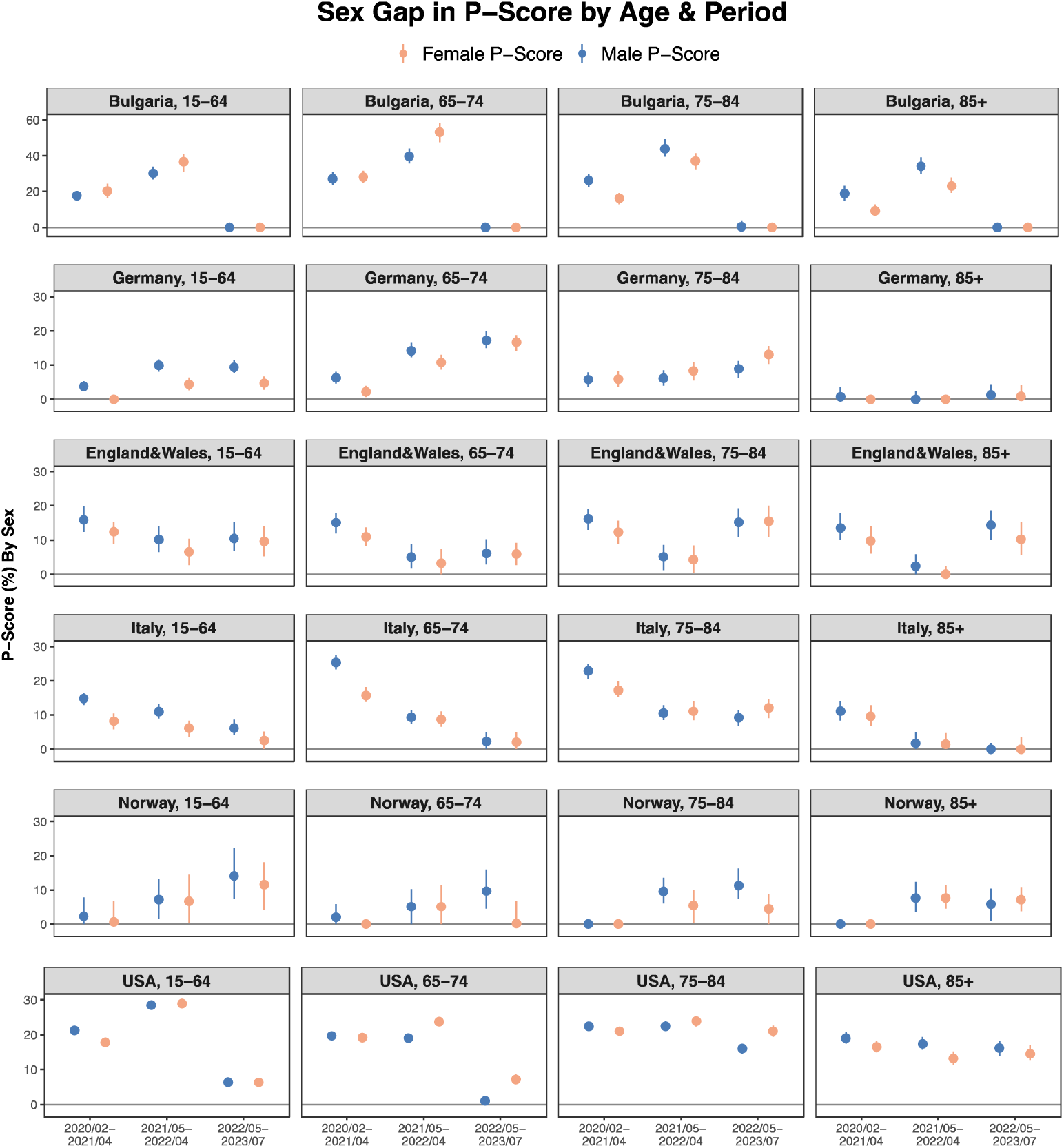
Excess death P-score by sex, period, age group and country. **Note:** the scale of the y-axis is fixed for all countries and age groups except Bulgaria Taken together, Figures 4 and 5 indicate that, although men were disproportionately more affected by the COVID-19 pandemic during the pre-vaccine phase in terms of absolute mortality, both sexes experienced proportionally similar increases in mortality when compared to pre-pandemic levels in most age groups and countries.

Figures S23 – S24 in the Supplementary Materials summarize the age-specific patterns in sex differences for excess mortality death rate (absolute gap), and P-score (relative gap) across three periods. Figure S23 illustrates the sex gaps as the arithmetic difference between male and female excess all-cause mortality death rate (per 100,000 person-years) by age group, period and country. During the pre-vaccine phase, a clear male disadvantage is evident in more than half of the countries, with the disadvantage increasing with age, where mortality rates are higher. In the post-vaccine and endemic phases, these differences in the absolute sex gap by age diminish.

The age pattern is less clear for relative sex gaps (male minus female P-score) (Figure S24). During the pre-vaccine period, males generally had higher P-Scores across most country-age combinations. However, in the post-vaccine and endemic periods, several age-country combinations show a shift towards a female disadvantage. In sum, a clear pattern of the male disadvantage increasing with age was only visible for excess mortality death rate (absolute gap) and mainly in the pre-vaccine period.

## Discussion

This study provides a comprehensive analysis of sex differences in excess mortality during the COVID-19 pandemic in 33 high-income countries, covering the pandemic’s progression through its pre-vaccine, post-vaccine, and endemic phases. Unlike earlier studies, we use an excess mortality framework, avoiding potential biases introduced by variability in testing and recording of COVID-19 deaths across countries. Our findings show that the COVID-19 pandemic temporarily amplified pre-existing sex differences in all-cause mortality.

Specifically, the observed male-to-female mortality ratios were higher than expected based on pre-pandemic trends, indicating a greater-than-usual male disadvantage during the pre-vaccine phase of the pandemic. In the endemic phase, observed ratios declined, becoming only slightly higher than expected—or, in some cases, even falling below them. This short-lived pattern of elevated sex differences in excess mortality contrasts with the 1918 influenza pandemic, which, had a longer lasting impact on sex differences in mortality. The 1918 flu caused unusually high male mortality, possibly due to co-infection with tuberculosis, which was more prevalent among men. Noymer and Garrene hypothesize that this the reduction in the sex gap in mortality in the US in the years following the pandemic was due to strong mortality selection that left behind cohorts of relatively healthier males ^33^. Our results suggest that COVID-19 is unlikely to produce similar long-term effects on sex differences in mortality in high-income countries.

Earlier studies consistently reported a male disadvantage in COVID-19 mortality ^19–21,23–30,32^. Our analysis of both absolute and relative sex gaps over time reveals a more nuanced picture. In absolute terms, and in line with earlier research, males experienced higher excess mortality during peak months, often exceeding pre-pandemic male-female mortality differences.

However, this was not a universal pattern; even in the pre-vaccine period, many age group–country combinations showed no significant male disadvantage. In contrast, relative measures showed less consistent male disadvantage. In several country–age combinations, females had a greater percentage increase in excess mortality relative to expected deaths, especially after the vaccine rollout. By the endemic phase, relative sex gaps had diminished across most countries and age groups. These reductions were often driven by steeper declines in male excess mortality, though in some cases, increased female mortality also contributed to the narrowing gap.

Comparing absolute vs. relative sex differences can highlight different aspects of changes in mortality. Absolute sex differences show the difference in the number of deaths during the crisis and how these differences change over time. A limitation of absolute sex differences is that men’s higher baseline mortality means a uniform increase in mortality risk leads to more deaths for men mathematically, saying little about whether men are more susceptible to the SARS-CoV-2 virus itself. Relative measures such as the P-score capture whether the percentage increase in mortality relative to expected baseline mortality is higher for one group or another, speaking more to whether sex differences in COVID-19 mortality are worse than for other causes of death. In some cases (e.g. in the post-vaccine phase in two younger age groups in Bulgaria), females saw a disproportionate increase in excess mortality compared to men.

Several potential mechanisms underlie the observed sex differences. Male disadvantage, particularly visible during the pre-vaccine phase of the pandemic likely resulted from a complex interplay between biological susceptibility to the virus ^9–17^ and gender-related social factors, such as health behaviours and occupational exposures. Women, on average, exhibit stronger and longer-lasting activation of key immune responses than men, and this response declines less with age in women. Women’s stronger innate and adaptive immune responses are influenced by hormonal factors, such as the immunomodulatory effects of estrogen, and genetic factors, including the higher expression of immune-related genes associated with the double X chromosome. These immune differences may help explain sex differences in COVID-19 outcomes ^15,16^.

Sex differences in excess mortality are also likely to be driven by social factors. Multiple surveys have shown that, on average, men were less likely to adhere to public health recommendations, such as mask-wearing, hand washing, and social distancing ^34,35^. On average, men reported a lower perceived risk of contracting COVIDJ19 and were less concerned about the potential health consequences of falling ill with the virus as compared to women ^50–52^.

Gender differences in structural exposure patterns may contribute to sex disparities in COVID-19 outcomes, but evidence on whether men faced higher exposure remains mixed. Previous studies have emphasised the strong connection between occupational roles and exposure to the COVID-19 virus ^53^. Essential workers, in particular, were at significantly higher risk of infection and mortality. These roles are often highly gender-segregated. In many high-income countries, women constitute a large share of frontline care, healthcare, and retail workers, which placed them at greater risk of exposure ^54–56^. By contrast, female-dominated sectors such as education and administrative roles were more likely to adopt remote work arrangements ^56,57^. Conversely, remote work was less feasible in male-dominated industries such as agriculture, security, logistics, construction, food production and manufacturing ^57^. These sectors reported some of the highest increases in mortality during the pandemic ^58^.

Overall, males experienced larger declines in excess mortality over the course of the pandemic, resulting in smaller absolute and relative sex differences over time. The diminished sex differences could be due to both biological and social factors, including higher levels of immunity due to vaccines and prior infections, mortality displacement, or changing preventive behaviours and risk of exposure. If males are more biologically susceptible to COVID-19 mortality, this risk would likely be highest in immune-naïve populations during the pre-vaccine stage of the pandemic. However, as immunity increased— either through vaccination or prior infection (for survivors)—this elevated risk likely diminished, narrowing sex differences. Additionally, vaccine hesitancy and uptake may have influenced these trends in a gendered way. Two nationally-representative surveys conducted in Europe and the US showed higher levels of COVID-19 vaccine hesitancy and lower uptake among women ^52,59^ This could help to explain more significant declines in male mortality in the later stages of the pandemic. By mid-2022, the highly transmissible Omicron variant led to widespread exposure, leaving very few immune-naïve individuals in most countries in the endemic phase. Over time, as COVID-19 becomes a less novel cause of death, we would expect sex differences in mortality to return closer to baseline levels.

Mortality displacement could also play a role: if more frail males died earlier in the pandemic compared to females, this could lead to reduced male mortality in later stages. Finally, occupational exposure-related factors, such as improved protective measures in male-dominated industries and other public health interventions, likely reduced mortality among initially more vulnerable males, narrowing or closing the sex gap in the post-vaccine and endemic phases of the pandemic.

Finally, women may have suffered more from the indirect effects of the pandemic, such as income and employment losses, disruption in healthcare access and gender-based violence ^60^, which potentially may have contributed to slower declines in excess mortality among women. These factors together could have led to larger reductions in absolute and relative mortality in males than in females, although further studies are needed to confirm these hypotheses.

We found large cross-country differences in the size and age patterns of both absolute and relative sex differences, suggesting the role of diverse social, economic, and healthcare factors in shaping sex-specific pandemic mortality trends. The largest absolute sex differences in excess mortality were observed in Eastern European countries, with small differences observed in Nordic countries, consistent with pre-pandemic sex differences ^61^. The pandemic exacerbated these pre-pandemic trends, further widening sex differences in regions where they were already high. Behavioural factors, such as high levels of smoking and chronic drinking among Eastern European men, factors known for increasing the risk of COVID-19 infection and severe disease progression ^62,63^, may have contributed to high excess mortality in this group. Smoking and drinking are also known risk factors for two diseases that contribute most to the pre-pandemic mortality sex gap, i.e. cardiovascular disease and lung cancer ^64,65^.

Gender occupational segregation may be another driver of cross-country mortality sex differences. The gender composition of occupations differs between countries in Europe, with some important regional differences ^66,67^. For example, Scandinavian countries experienced rapid occupational gender desegregation in recent decades, whereas segregation increased in Mediterranean countries and Eastern Europe ^66^. This could explain why sex differences reported in our study were, on average, larger in Eastern Europe and Mediterranean countries than in Scandinavia.

Sex differences in COVID-19-associated mortality reflect a complex interplay of genetic, biological, and social factors. Cross-country variations in the magnitude of these differences highlight the significance of social determinants as key drivers of sex differences in mortality. The widespread male disadvantage in absolute COVID-related mortality, particularly evident during the pre-vaccine stage, likely underscores males’ less effective immune response and vulnerability to novel pathogens, consistent with greater male vulnerability during previous health crises ^8^. However, the changing nature of sex-specific mortality differences and the contrasting patterns observed in absolute versus relative indicators suggests greater nuance. Specifically, situating COVID-19 mortality within pre-existing sex differences in mortality reveals that from a relative perspective, the pandemic’s effects were often comparable between sexes and, in some cases, worse for women.

### Limitations

Our analysis has limitations. First, our conclusions are limited to the selected high-income countries with efficient vital registration systems. Second, our population counts for 2022-2023 were extrapolated from 2021. This may lead to bias due to not accounting for pandemic deaths and migration. Third, the STMF data is available only for very broad age groups. For this reason, we were able to only roughly account for between-country differences in age structure^68^. Finally, our data does not account for gender identity and is available only for people categorised as women and men.

### Conclusion

We present a comprehensive analysis of sex differences in excess mortality during the COVID-19 pandemic, covering 33 high-income countries across three phases: pre-vaccine, post-vaccine, and endemic. Addressing limitations of earlier studies, we use an excess mortality framework and examine both absolute and relative measures of sex differences, allowing for more reliable comparisons across time and place. While the prevailing narrative highlights a persistent male disadvantage in COVID-19 mortality, our findings show that this pattern was context-specific. Male disadvantage was evident during high-mortality months in the pre-vaccine phase and in absolute terms, but it was not universal. When using relative measures, which account for baseline mortality differences, the male disadvantage largely disappeared—or even reversed—in most age group–country combinations. These findings underscore the importance of using both absolute and relative metrics to understand inequalities in mortality and suggest that COVID-19 did not produce lasting shifts in sex differences in mortality.

## Competing interest statement

None declared.

## Funding

This work was supported by the European Research Council (ERC-2021-CoG-101002587) and the Leverhulme Trust (Grant RC-2018-003) for the Leverhulme Centre for Demographic Science. The researchers involved in this study declare full independence from the funders. The funding bodies had no role in the design of the study, analysis, or interpretation of the data; the writing of the manuscript; or the decision to submit it for publication.

## Transparency declaration

The lead author affirms that the manuscript is an honest, accurate, and transparent account of the study being reported; that no important aspects of the study have been omitted; and that any discrepancies from the study as planned (and, if relevant, registered) have been explained.

## Ethics approval

This research project does not require ethics approval as it uses only national level vital statistics that are publicly available online.

## Data sharing

Data used in the analyses of this manuscript is publicly available at https://www.mortality.org/Data/STMF. Metadata and code will be available at peer review and upon publication.

## Data Availability

Materials for reproduction, including metadata and scripts, will be available at peer review and upon publication.

## Notes

### Competing Interest Statement

The authors have declared no competing interest.

### Author Declarations

This study uses ONLY publicly available, anonymised, aggregate data on all-cause mortality from the Short-Term Mortality Fluctuations (STMF) database, which does not include individual-level or identifiable information. As such, it does not require ethics approval under our institution's guidelines. Data link: www.mortality.org/Data/STMF

