## Supplementary Information for "Sex Differences in Excess Mortality During the COVID-19 Pandemic"

Materials

[Figure S 1. Monthly trends in excess mortality death rate (per 100,000 person-years) by sex, age, and country [Austria, Belgium, Croatia, Czechia, Denmark, and Hungary]. Part 1 of 5..	3](#_Toc202736887)

[Figure S 2. Monthly trends in excess mortality death rate (per 100,000 person-years) by sex, age, and country [Estonia, Finland, France, Greece, Iceland, and Israel]. Part 2 of 5.	4](#_Toc202736888)

[Figure S 3. Monthly trends in excess mortality death rate (per 100,000 person-years) by sex, age, and country [Latvia, Lithuania, Luxembourg, Netherlands, New Zealand, and Northern Ireland]. Part 3 of 5.	5](#_Toc202736889)

[Figure S 4. Monthly trends in excess mortality death rate (per 100,000 person-years) by sex, age, and country [Poland, Portugal, Scotland, Slovakia, Slovenia and South Korea]. Part 4 of 5.	6](#_Toc202736890)

[Figure S 5. Monthly trends in excess mortality death rate (per 100,000 person-years) by sex, age, and country [Spain, Sweden, and Switzerland]. Part 5 of 5.	7](#_Toc202736891)

[Figure S 6. Monthly trends in excess death P-scores by sex, age and country [Austria, Belgium, Croatia, Czechia, Denmark, and Hungary]. Part 1 of 5.	8](#_Toc202736892)

[Figure S 7. Monthly trends in excess death P-scores by sex, age and country [Estonia, Finland, France, Greece, Iceland, and Israel]. Part 2 of 5.	9](#_Toc202736893)

[Figure S 8. Monthly trends in excess death P-scores by sex, age and country [Latvia, Lithuania, Luxembourg, Netherlands, New Zealand, and Northern Ireland]. Part 3 of 5.	10](#_Toc202736894)

[Figure S 9. Monthly trends in excess death P-scores by sex, age and country [Poland, Portugal, Scotland, Slovakia, Slovenia and South Korea]. Part 4 of 5.	11](#_Toc202736895)

[Figure S 10. Monthly trends in excess death P-scores by sex, age and country [Spain, Sweden, and Switzerland]. Part 5 of 5.	12](#_Toc202736896)

[Figure S 11. All-cause excess mortality rate by sex (left y-axis) and the sex gap in excess mortality (male minus female, right y-axis), shown by age group, period, and country [Bulgaria, England&Wales, Germany, Italy, Norway, and USA]. Part 1 of 6.	13](#_Toc202736897)

[Figure S 12. All-cause excess mortality rate by sex (left y-axis) and the sex gap in excess mortality (male minus female, right y-axis), shown by age group, period, and country [Austria, Belgium, Croatia, Czechia, Denmark, and Hungary]. Part 2 of 6.	14](#_Toc202736898)

[Figure S 13. All-cause excess mortality rate by sex (left y-axis) and the sex gap in excess mortality (male minus female, right y-axis), shown by age group, period, and country [Estonia, Finland, France, Greece, Iceland, and Israel]. Part 3 of 6.	15](#_Toc202736899)

[Figure S 14. All-cause excess mortality rate by sex (left y-axis) and the sex gap in excess mortality (male minus female, right y-axis), shown by age group, period, and country [Latvia, Lithuania, Luxembourg, Netherlands, New Zealand, and Northern Ireland]. Part 4 of 6.	16](#_Toc202736900)

[Figure S 15. All-cause excess mortality rate by sex (left y-axis) and the sex gap in excess mortality (male minus female, right y-axis), shown by age group, period, and country [Poland, Portugal, Scotland, Slovakia, Slovenia and South Korea]. Part 5 of 6.	17](#_Toc202736901)

[Figure S 16. All-cause excess mortality rate by sex (left y-axis) and the sex gap in excess mortality (male minus female, right y-axis), shown by age group, period, and country [Spain, Sweden, and Switzerland]. Part 6 of 6.	18](#_Toc202736902)

[Figure S 17. Excess death P-score by sex (left y-axis) and the sex gap in P-score (male minus female, right y-axis), shown by age group, period, and country [Bulgaria, England&Wales, Germany, Italy, Norway, USA]. Part 1 of 6.	19](#_Toc202736903)

[Figure S 18. Excess death P-score by sex (left y-axis) and the sex gap in P-score (male minus female, right y-axis), shown by age group, period, and country [Austria, Belgium, Croatia, Czechia, Denmark, and Hungary]. Part 2 of 6.	20](#_Toc202736904)

[Figure S 19. Excess death P-score by sex (left y-axis) and the sex gap in P-score (male minus female, right y-axis), shown by age group, period, and country [Estonia, Finland, France, Greece, Iceland, Israel]. Part 3 of 6.	21](#_Toc202736905)

[Figure S 20. Excess death P-score by sex (left y-axis) and the sex gap in P-score (male minus female, right y-axis), shown by age group, period, and country [Latvia, Lithuania, Luxembourg, Netherlands, New Zealand, Northern Ireland]. Part 4 of 6.	22](#_Toc202736906)

[Figure S 21. Excess death P-score by sex (left y-axis) and the sex gap in P-score (male minus female, right y-axis), shown by age group, period, and country [Poland, Portugal, Scotland, Slovakia, Slovenia and South Korea]. Part 5 of 6.	23](#_Toc202736907)

[Figure S 22. Excess death P-score by sex (left y-axis) and the sex gap in P-score (male minus female, right y-axis), shown by age group, period, and country [Spain, Sweden, and Switzerland]. Part 6 of 6.	24](#_Toc202736908)

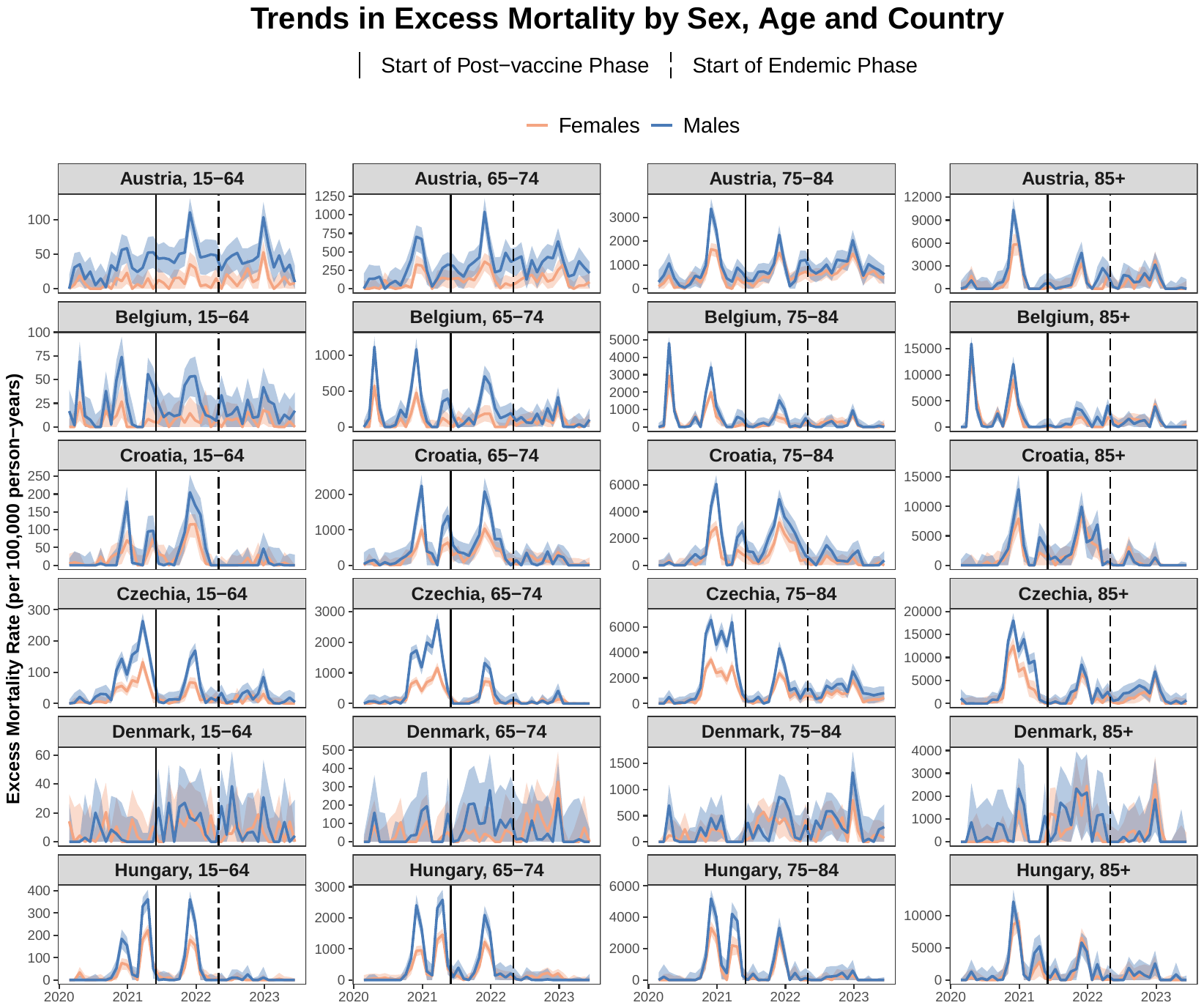

Figure S 1. Monthly trends in excess mortality death rate (per 100,000 person-years) by sex, age, and country [Austria, Belgium, Croatia, Czechia, Denmark, and Hungary]. Part 1 of 5..

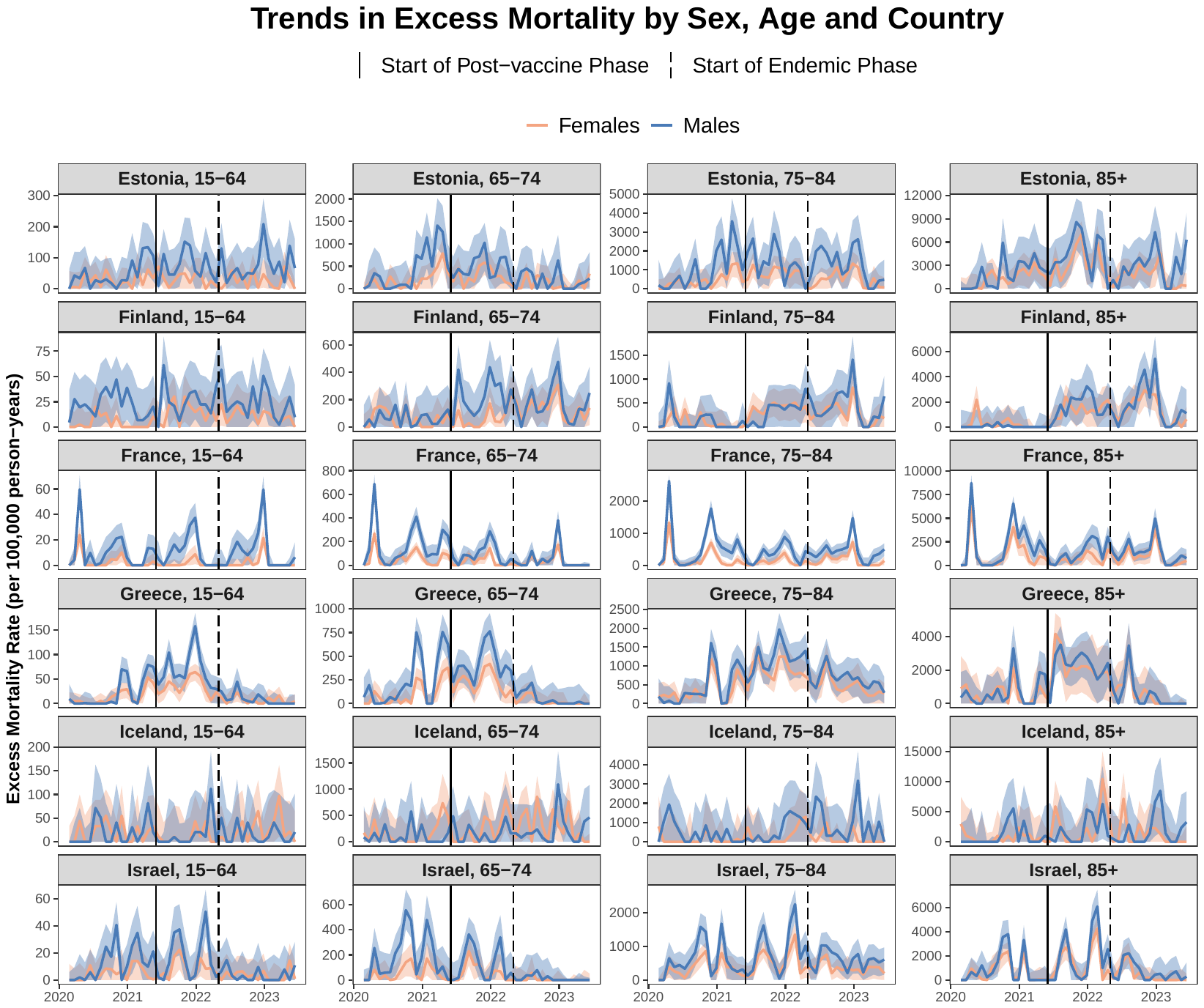

Figure S 2. Monthly trends in excess mortality death rate (per 100,000 person-years) by sex, age, and country [Estonia, Finland, France, Greece, Iceland, and Israel]. Part 2 of 5.

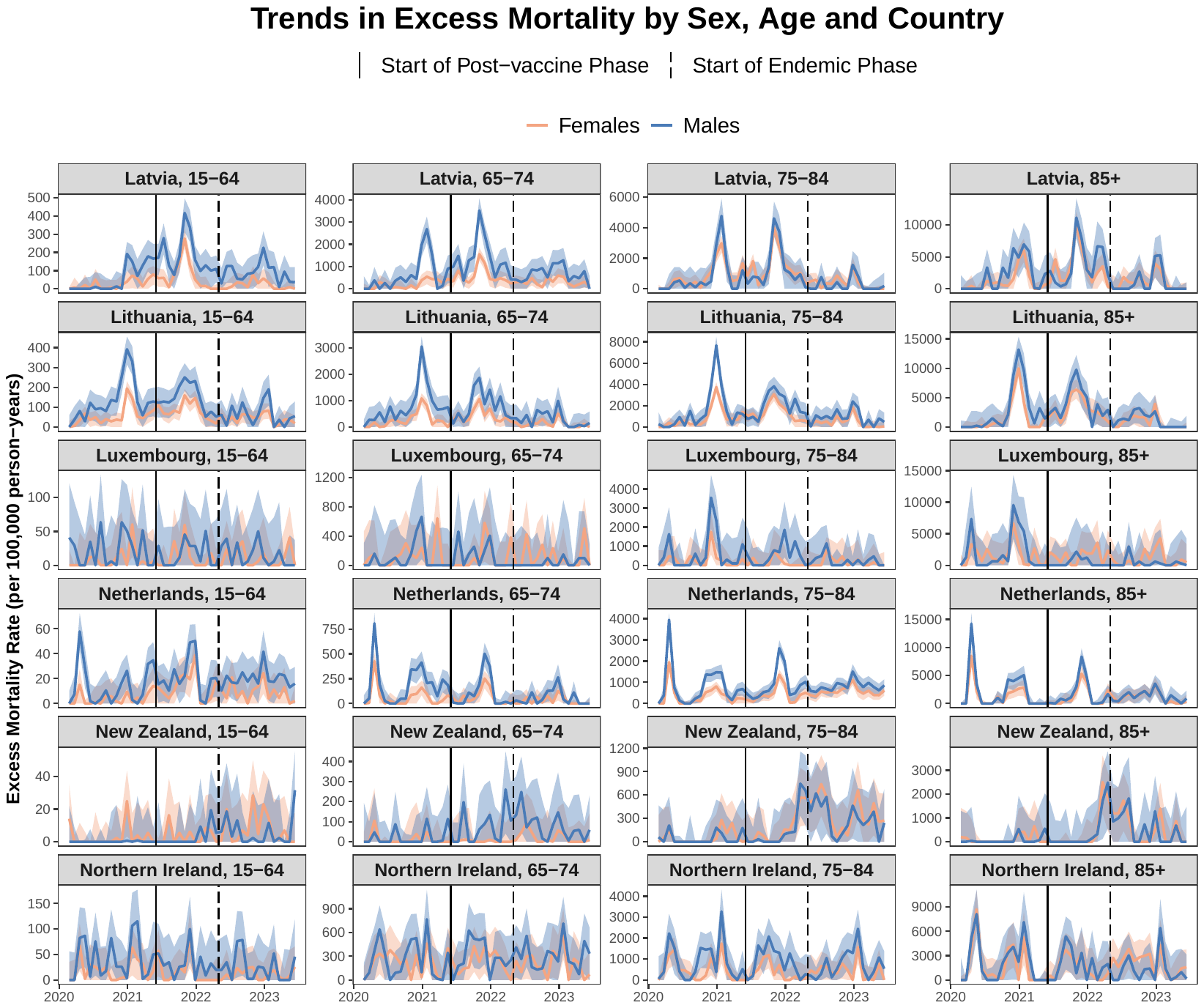

Figure S 3. Monthly trends in excess mortality death rate (per 100,000 person-years) by sex, age, and country [Latvia, Lithuania, Luxembourg, Netherlands, New Zealand, and Northern Ireland]. Part 3 of 5.

Figure S1. (continued) Monthly trends in excess mortality death rate (per 100,000 person-years) by sex, age, and country.

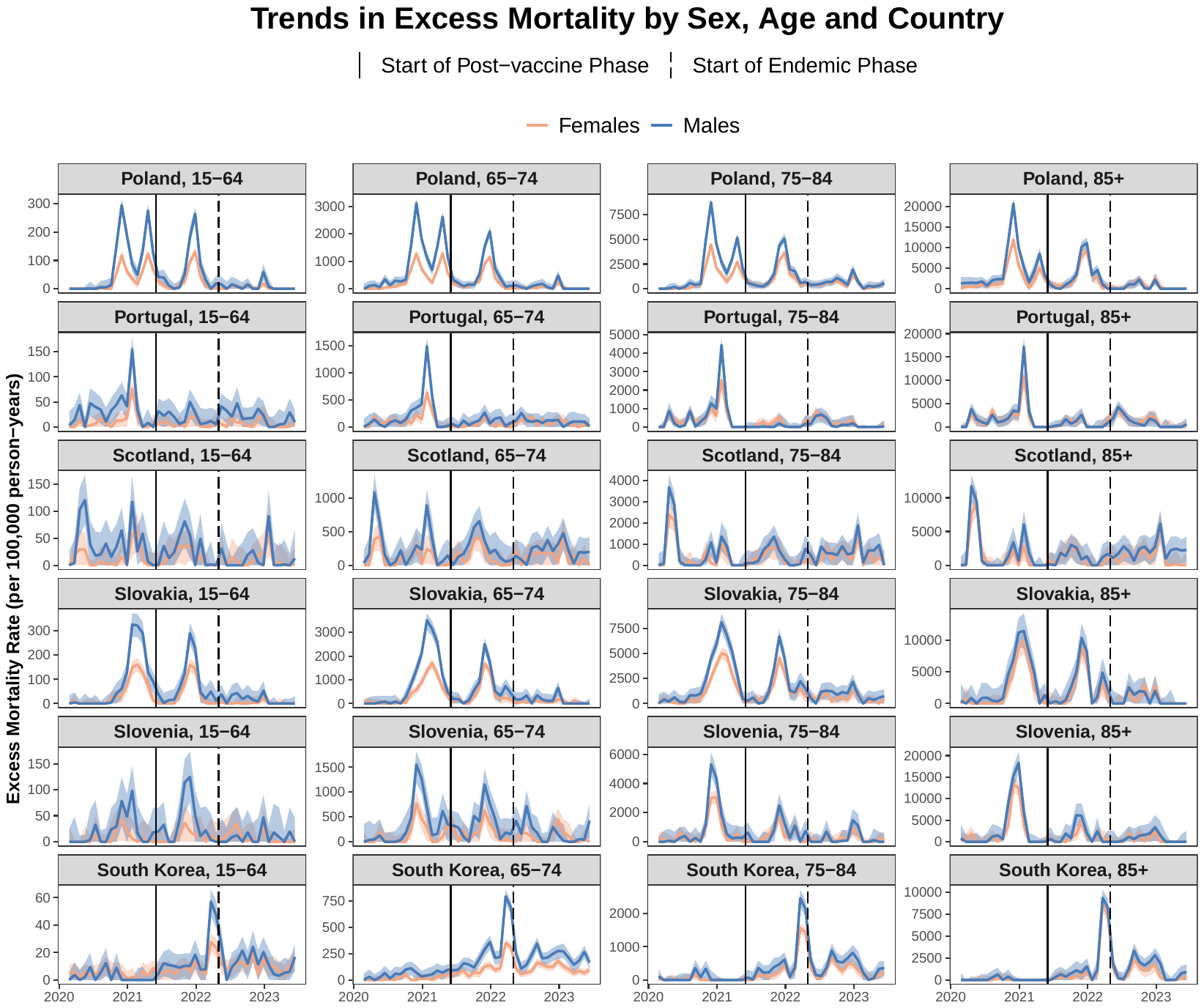

Figure S 4. Monthly trends in excess mortality death rate (per 100,000 person-years) by sex, age, and country [Poland, Portugal, Scotland, Slovakia, Slovenia and South Korea]. Part 4 of 5.

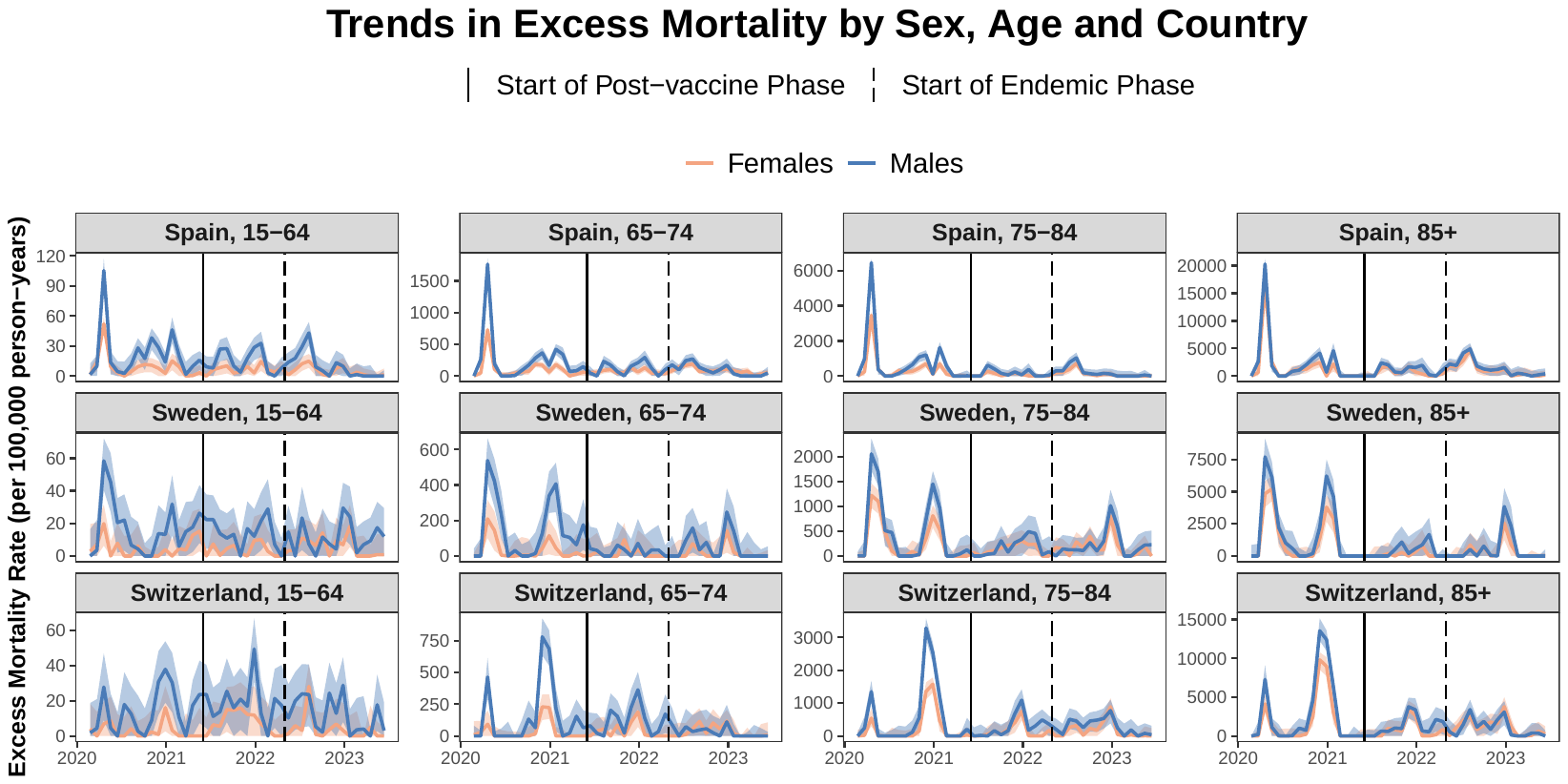

Figure S 5. Monthly trends in excess mortality death rate (per 100,000 person-years) by sex, age, and country [Spain, Sweden, and Switzerland]. Part 5 of 5.

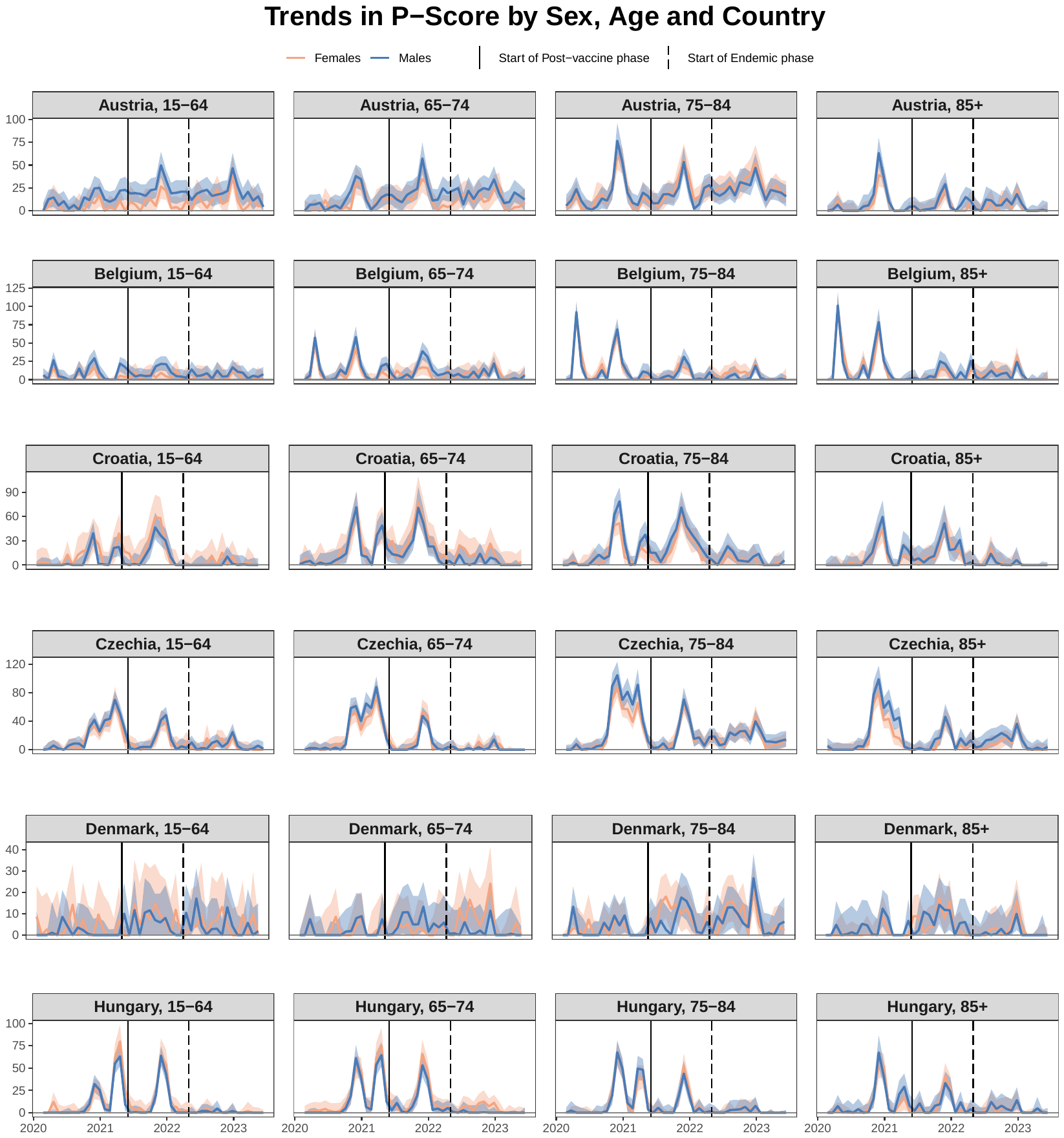

Figure S 6. Monthly trends in excess death P-scores by sex, age and country [Austria, Belgium, Croatia, Czechia, Denmark, and Hungary]. Part 1 of 5.

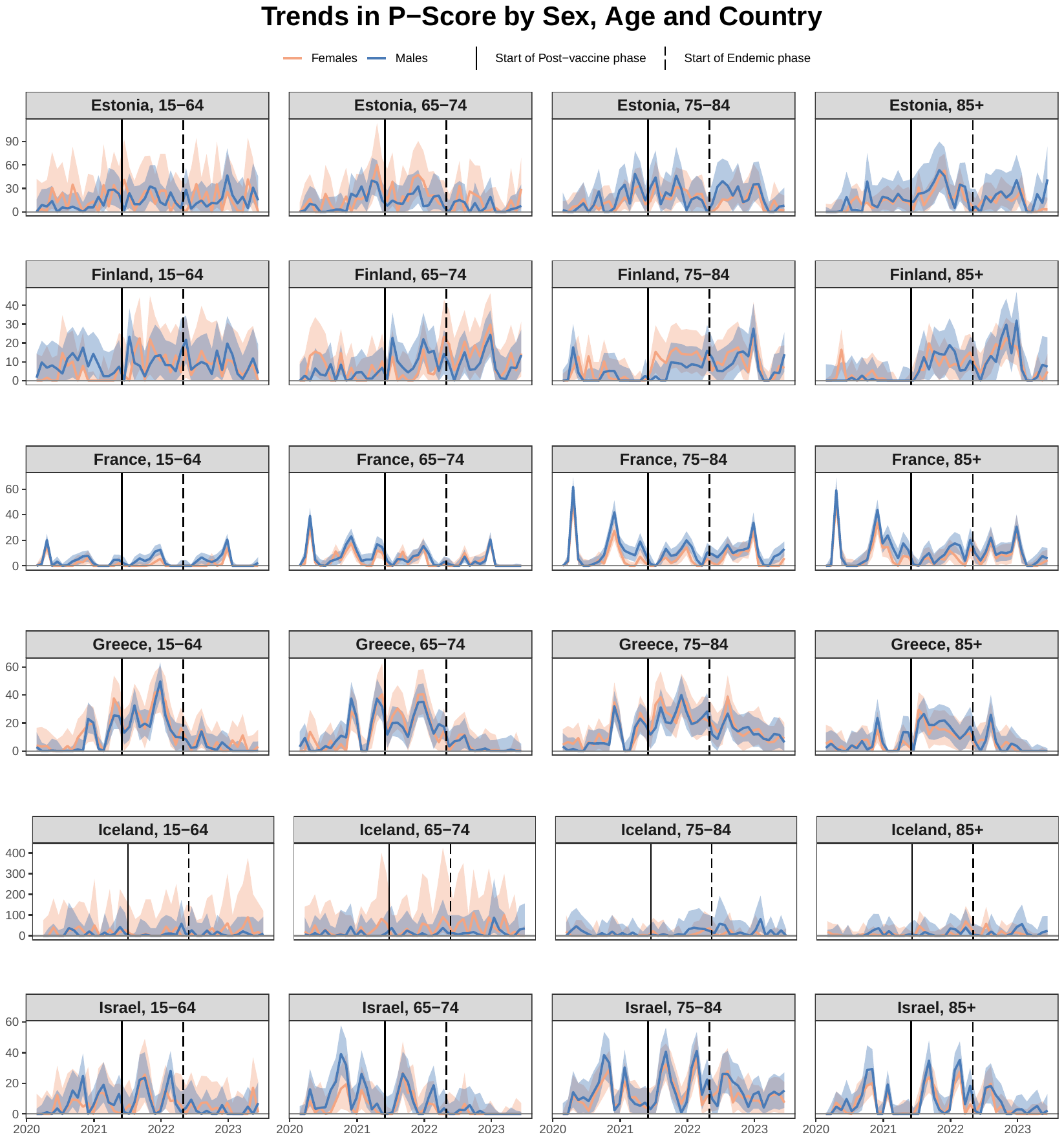

Figure S 7. Monthly trends in excess death P-scores by sex, age and country [Estonia, Finland, France, Greece, Iceland, and Israel]. Part 2 of 5.

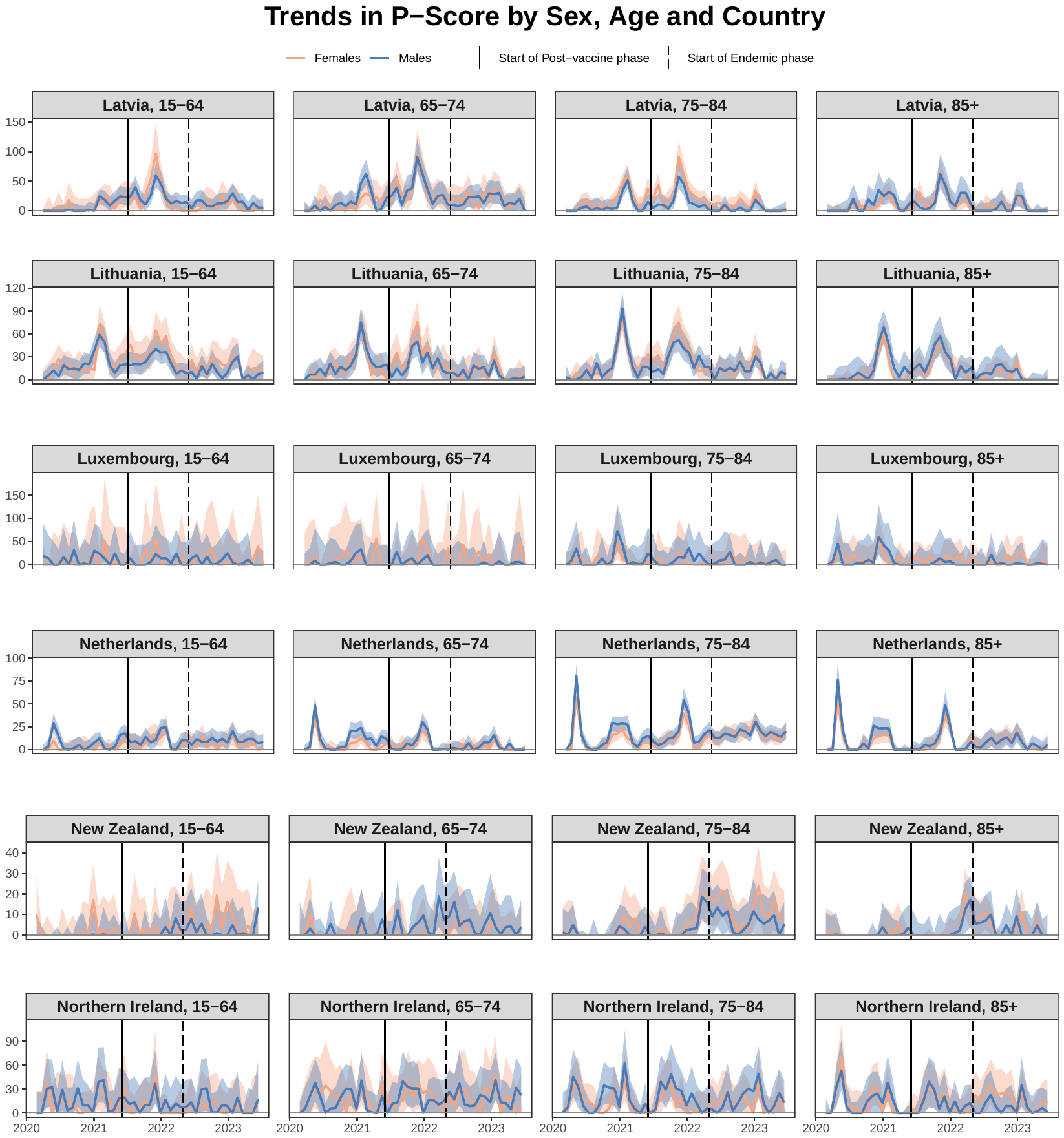

Figure S 8. Monthly trends in excess death P-scores by sex, age and country [Latvia, Lithuania, Luxembourg, Netherlands, New Zealand, and Northern Ireland]. Part 3 of 5.

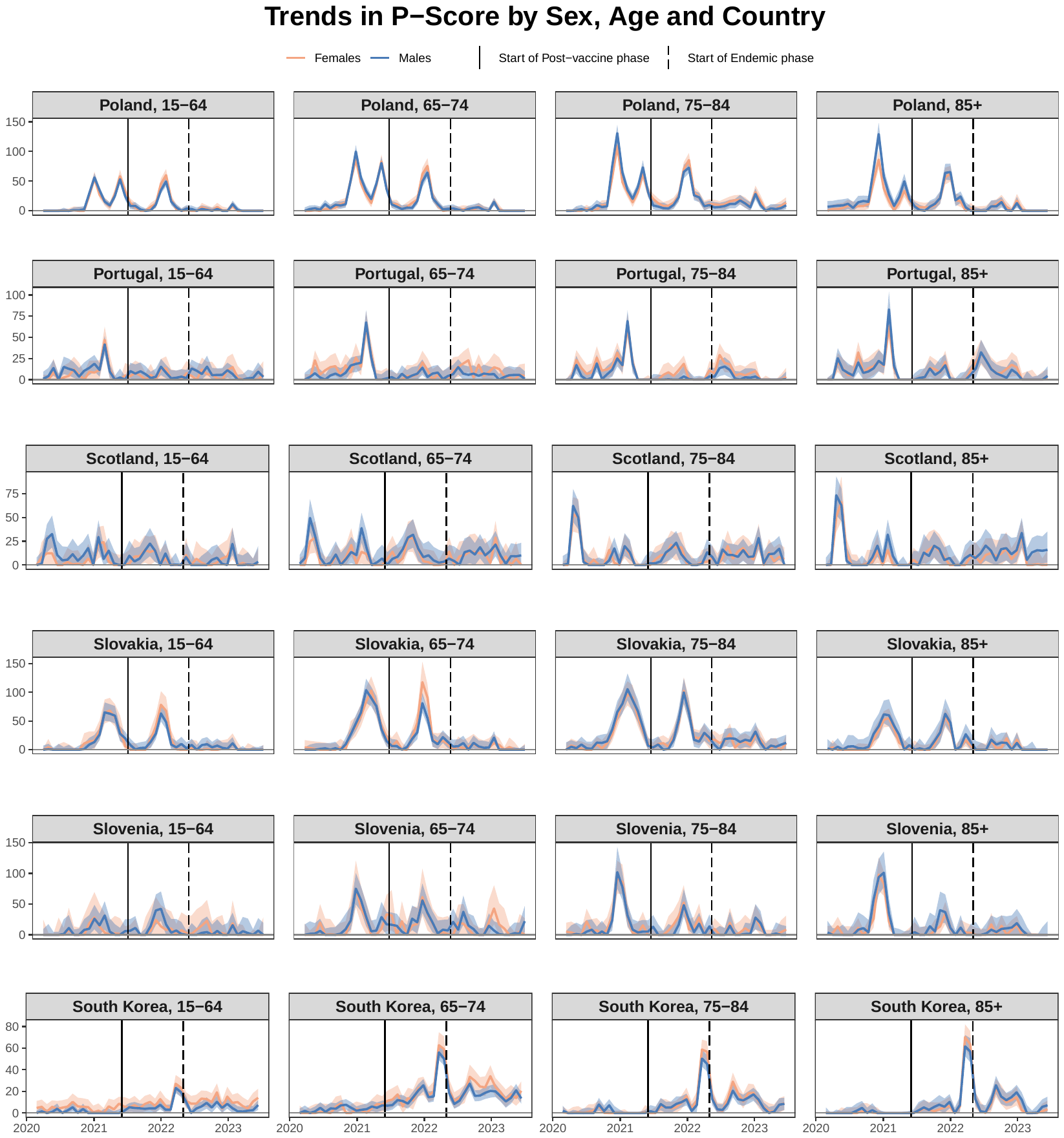

Figure S 9. Monthly trends in excess death P-scores by sex, age and country [Poland, Portugal, Scotland, Slovakia, Slovenia and South Korea]. Part 4 of 5.

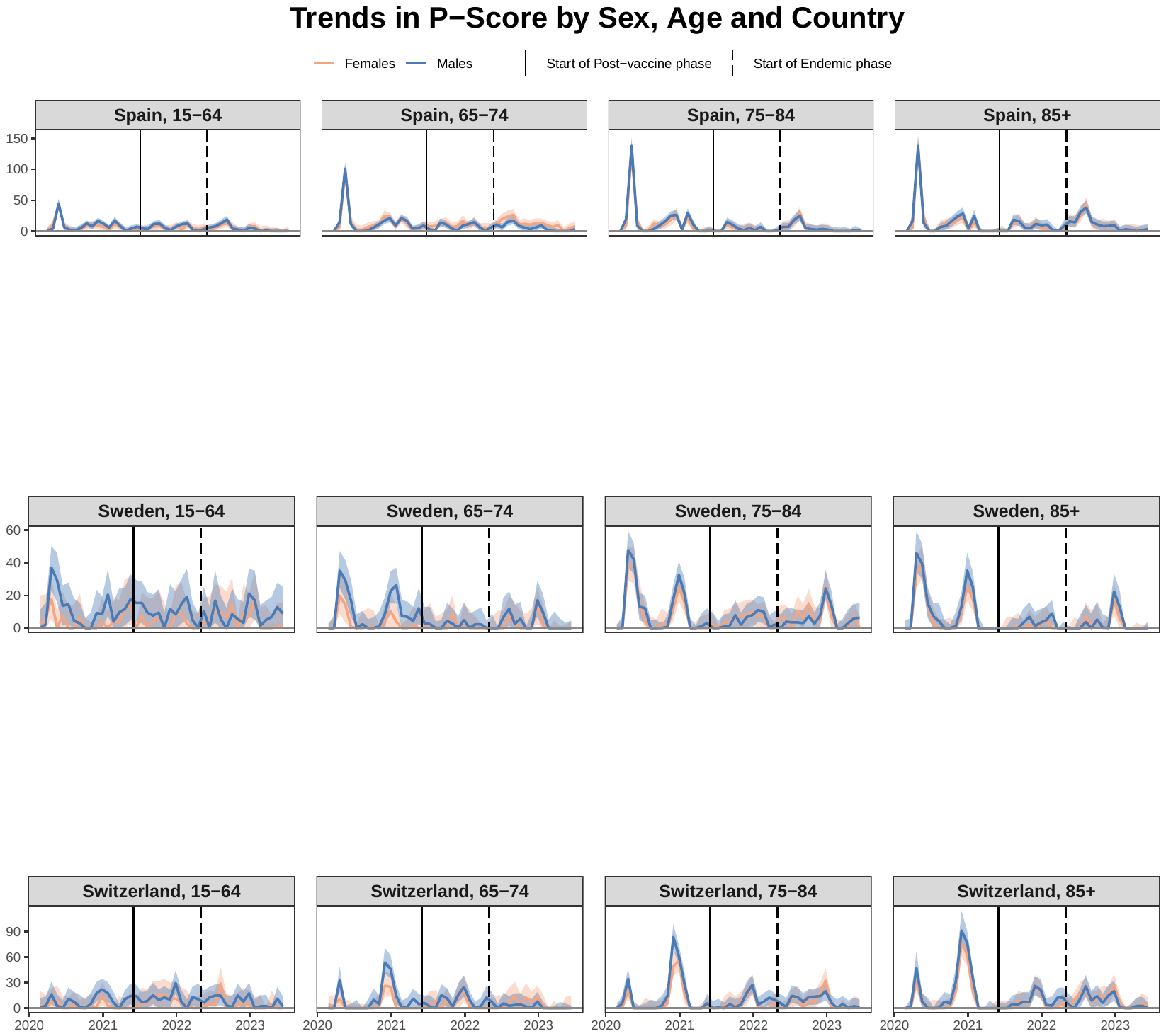

Figure S 10. Monthly trends in excess death P-scores by sex, age and country [Spain, Sweden, and Switzerland]. Part 5 of 5.

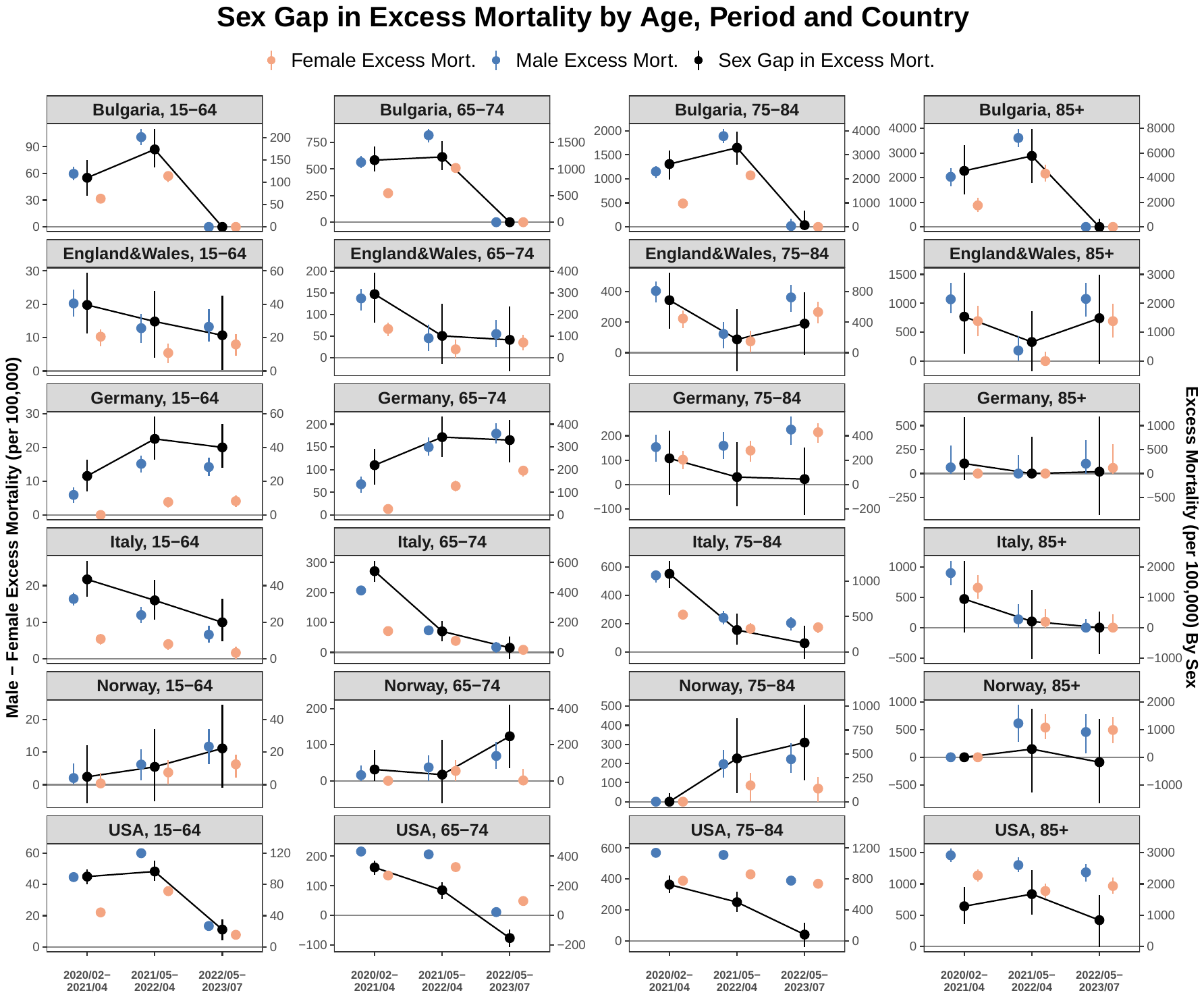

Figure S 11. All-cause excess mortality rate by sex (left y-axis) and the sex gap in excess mortality (male minus female, right y-axis), shown by age group, period, and country [Bulgaria, England&Wales, Germany, Italy, Norway, and USA]. Part 1 of 6.

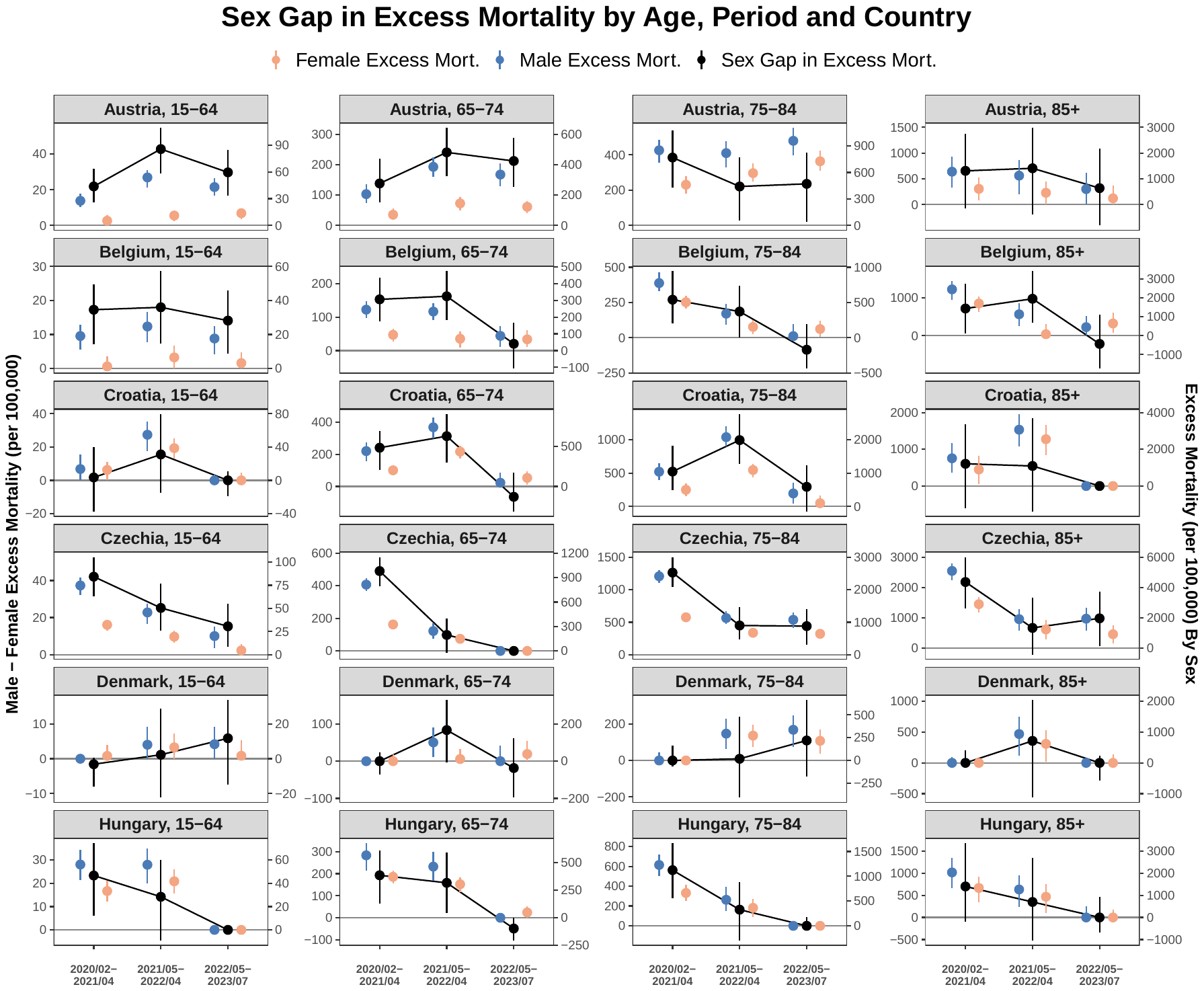

Figure S 12. All-cause excess mortality rate by sex (left y-axis) and the sex gap in excess mortality (male minus female, right y-axis), shown by age group, period, and country [Austria, Belgium, Croatia, Czechia, Denmark, and Hungary]. Part 2 of 6.

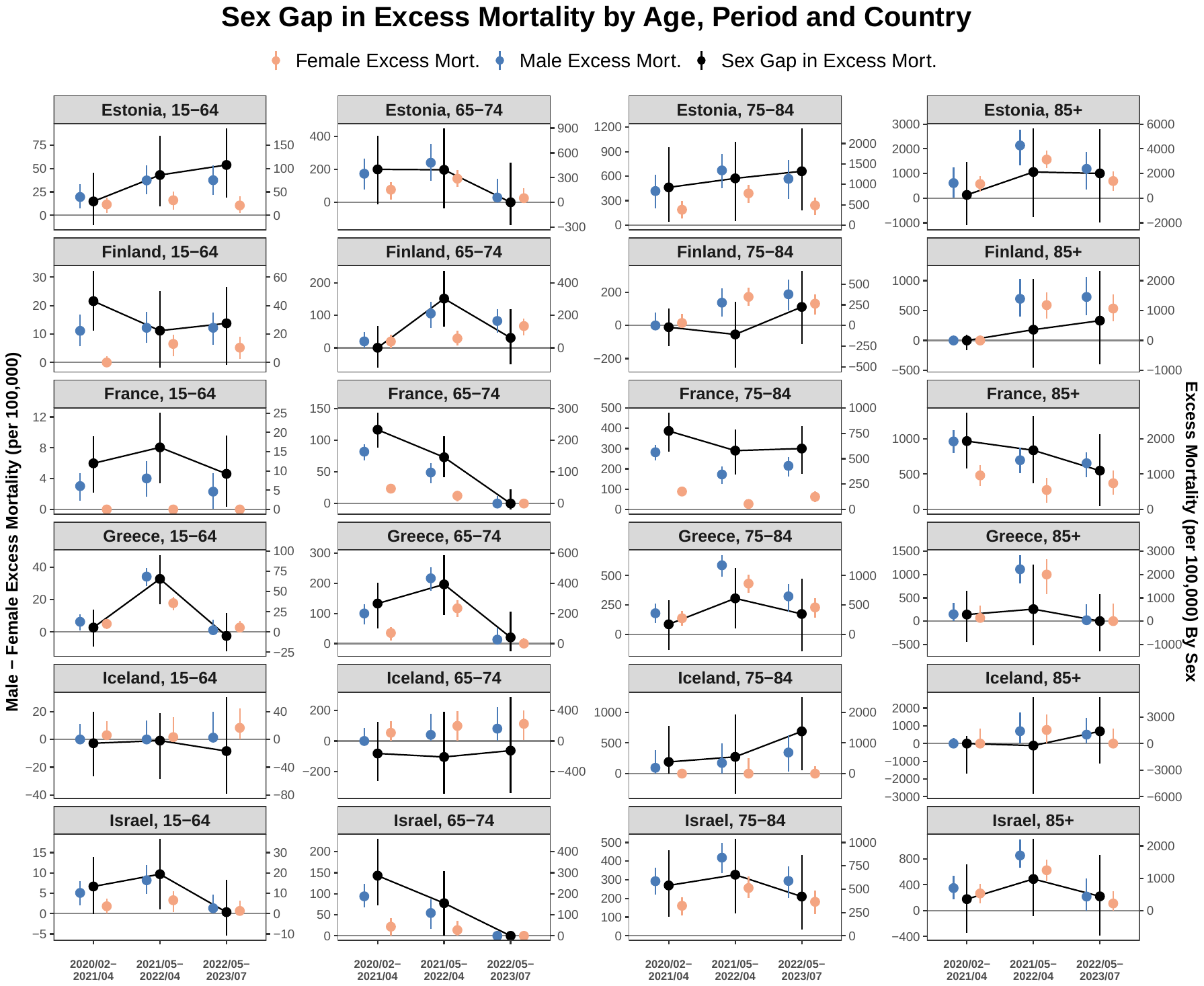

Figure S 13. All-cause excess mortality rate by sex (left y-axis) and the sex gap in excess mortality (male minus female, right y-axis), shown by age group, period, and country [Estonia, Finland, France, Greece, Iceland, and Israel]. Part 3 of 6.

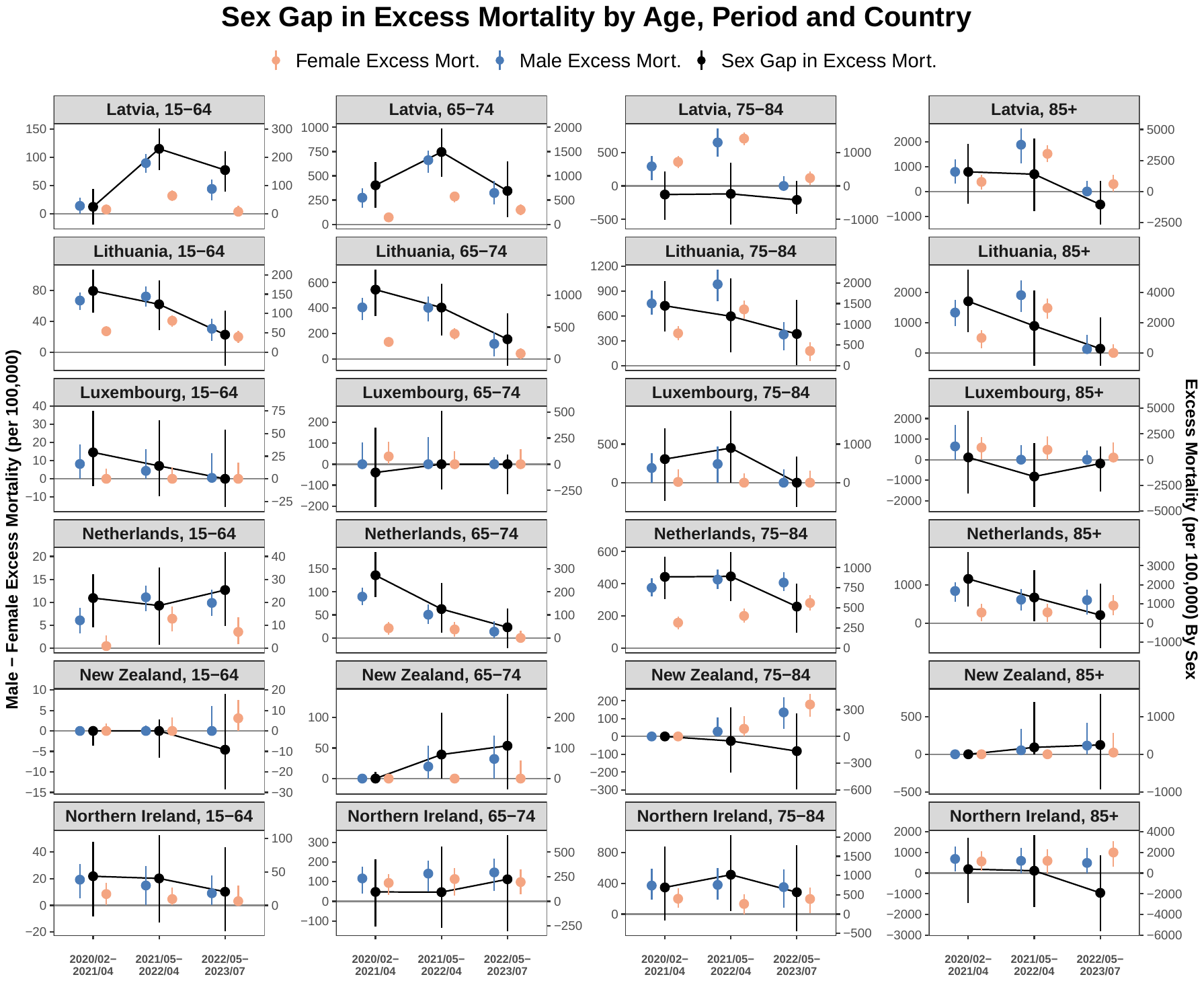

Figure S 14. All-cause excess mortality rate by sex (left y-axis) and the sex gap in excess mortality (male minus female, right y-axis), shown by age group, period, and country [Latvia, Lithuania, Luxembourg, Netherlands, New Zealand, and Northern Ireland]. Part 4 of 6.

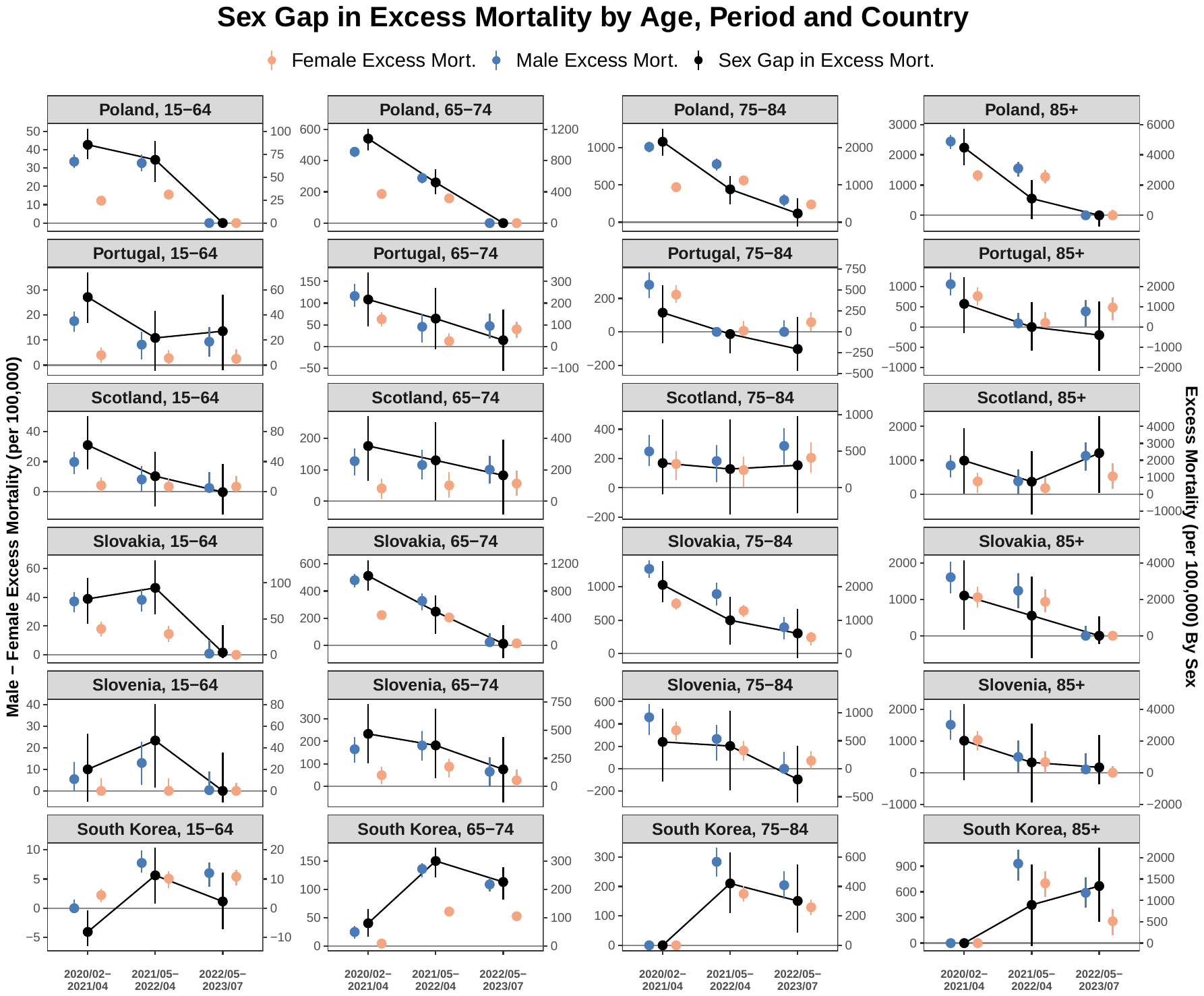

Figure S 15. All-cause excess mortality rate by sex (left y-axis) and the sex gap in excess mortality (male minus female, right y-axis), shown by age group, period, and country [Poland, Portugal, Scotland, Slovakia, Slovenia and South Korea]. Part 5 of 6.

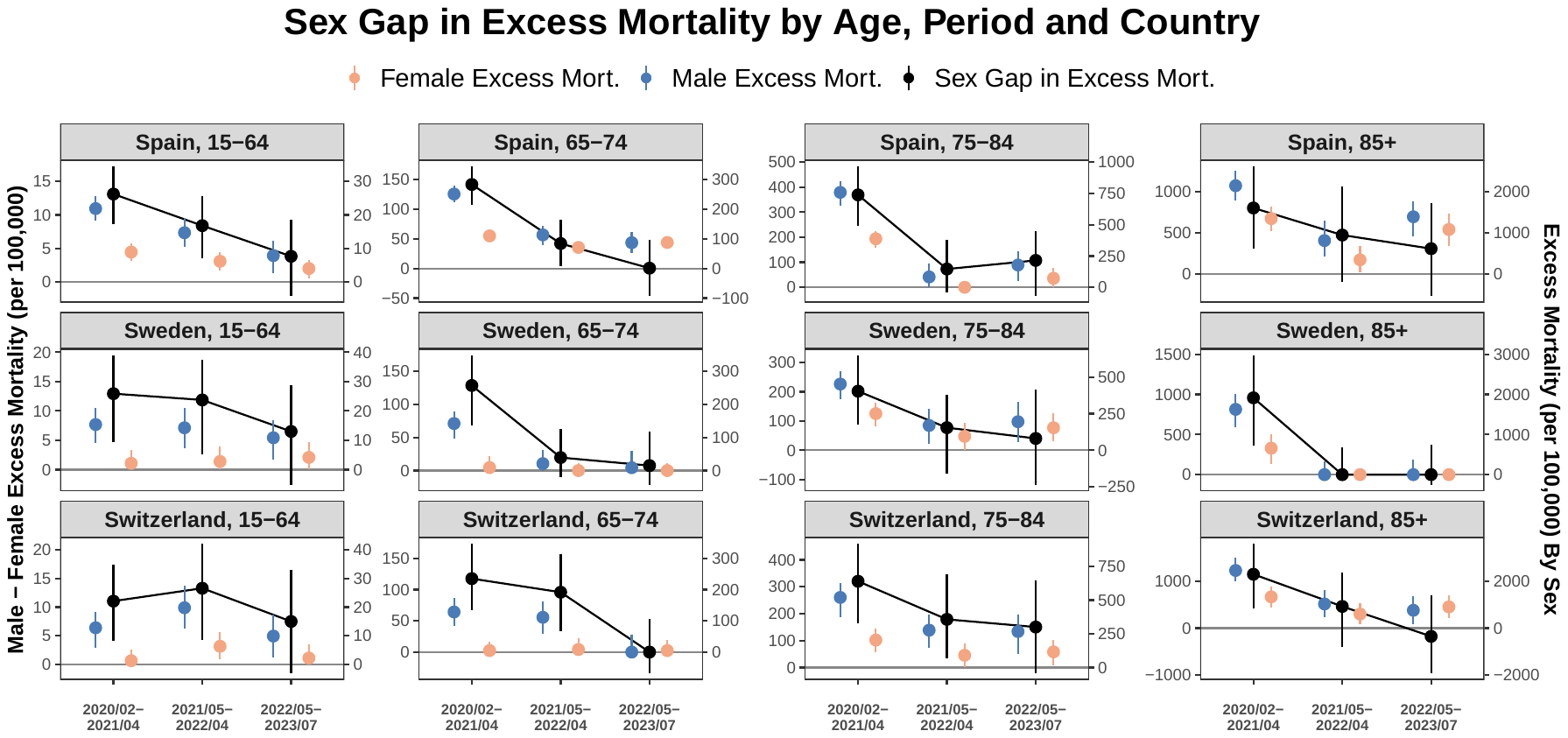

Figure S 16. All-cause excess mortality rate by sex (left y-axis) and the sex gap in excess mortality (male minus female, right y-axis), shown by age group, period, and country [Spain, Sweden, and Switzerland]. Part 6 of 6.

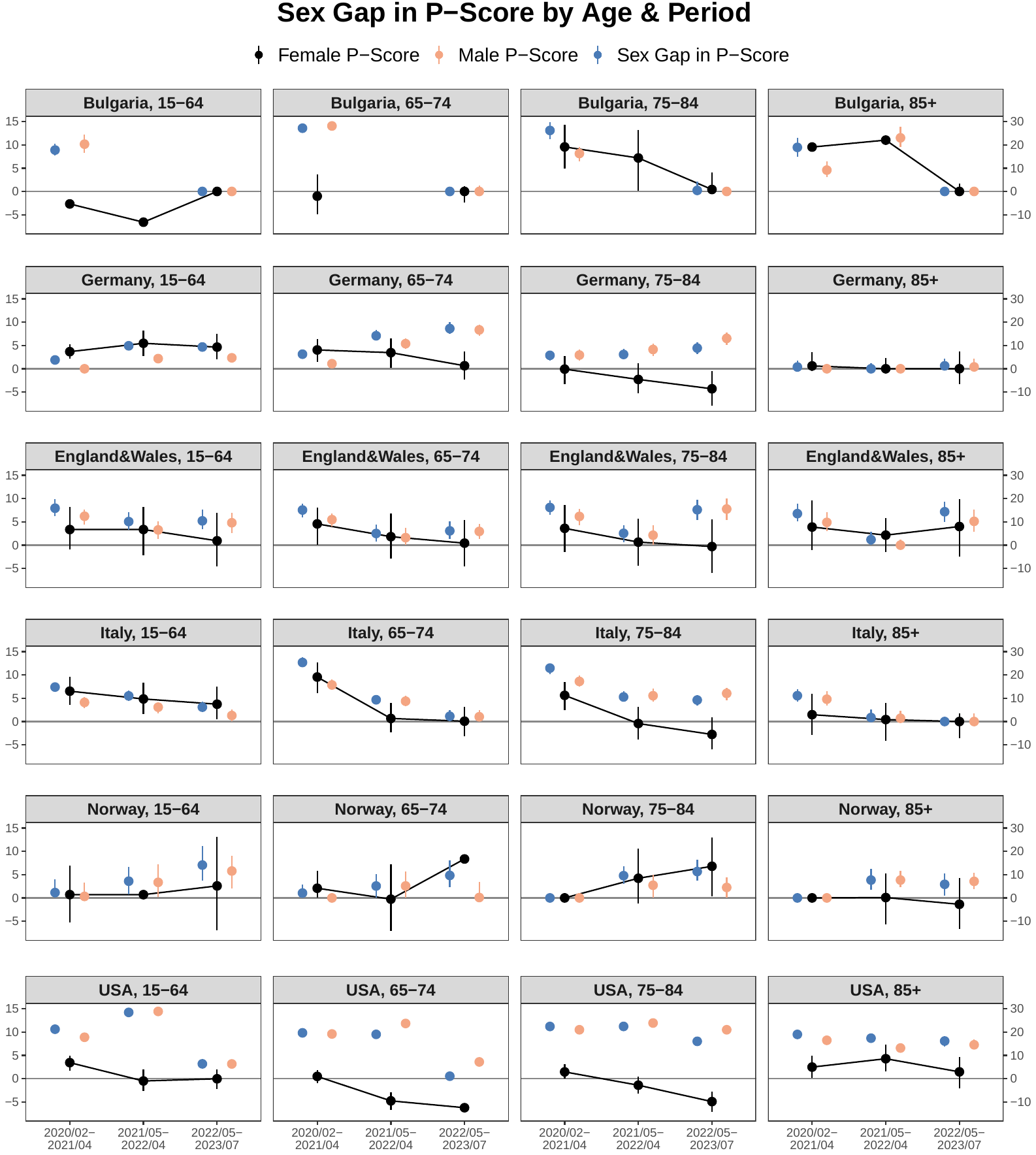

Figure S 17. Excess death P-score by sex (left y-axis) and the sex gap in P-score (male minus female, right y-axis), shown by age group, period, and country [Bulgaria, England&Wales, Germany, Italy, Norway, USA]. Part 1 of 6.

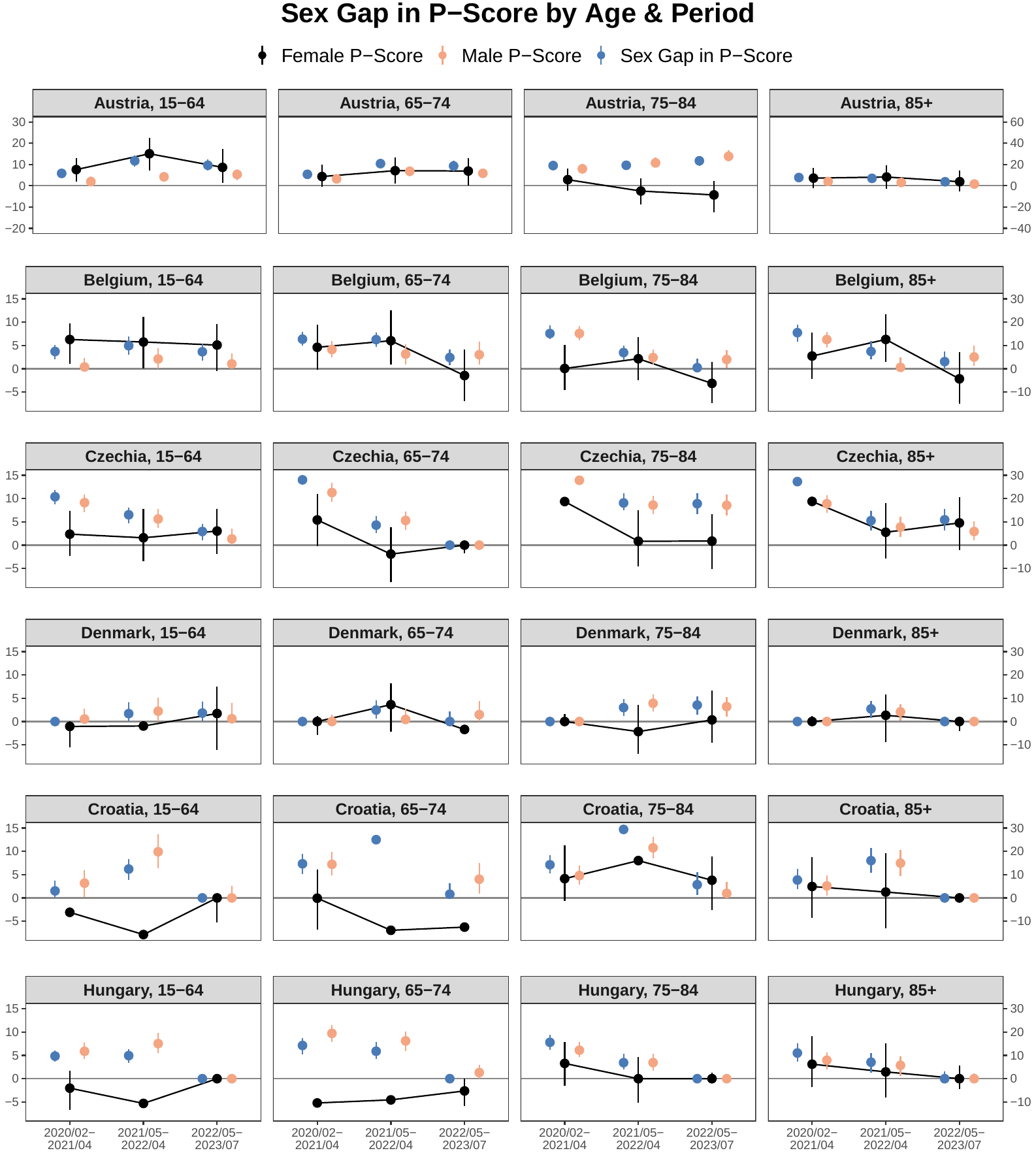

Figure S 18. Excess death P-score by sex (left y-axis) and the sex gap in P-score (male minus female, right y-axis), shown by age group, period, and country [Austria, Belgium, Croatia, Czechia, Denmark, and Hungary]. Part 2 of 6.

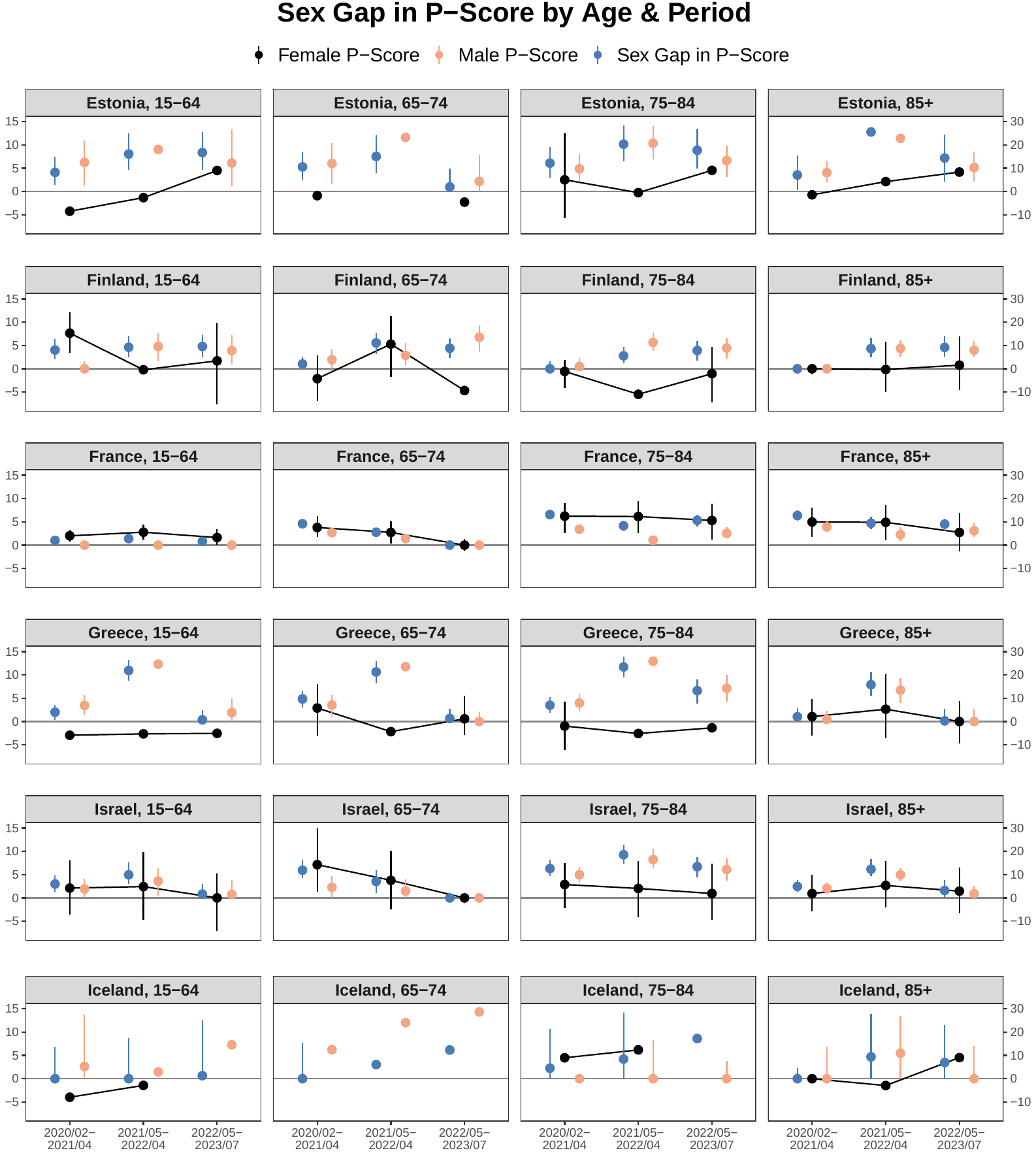

Figure S 19. Excess death P-score by sex (left y-axis) and the sex gap in P-score (male minus female, right y-axis), shown by age group, period, and country [Estonia, Finland, France, Greece, Iceland, Israel]. Part 3 of 6.

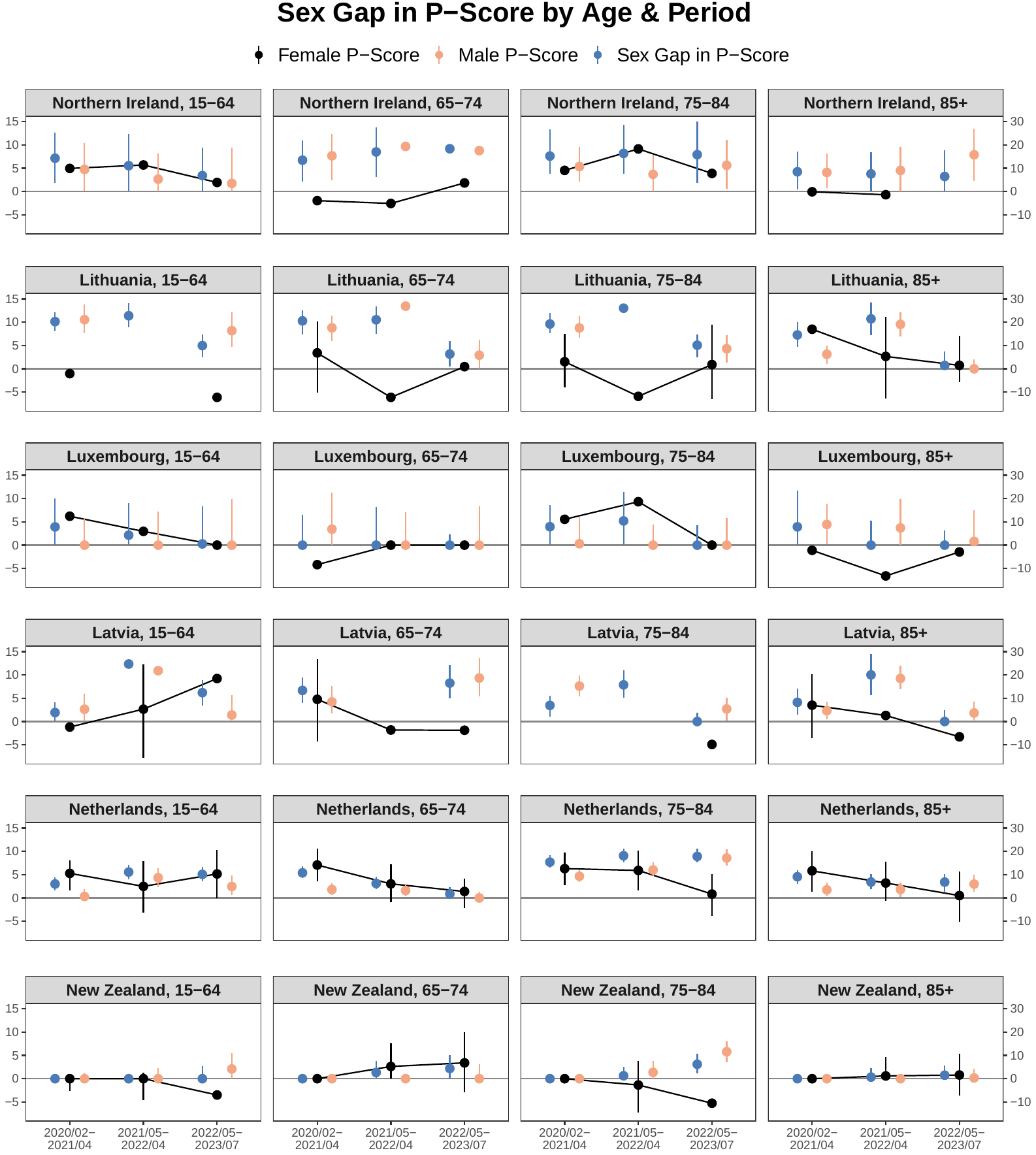

Figure S 20. Excess death P-score by sex (left y-axis) and the sex gap in P-score (male minus female, right y-axis), shown by age group, period, and country [Latvia, Lithuania, Luxembourg, Netherlands, New Zealand, Northern Ireland]. Part 4 of 6.

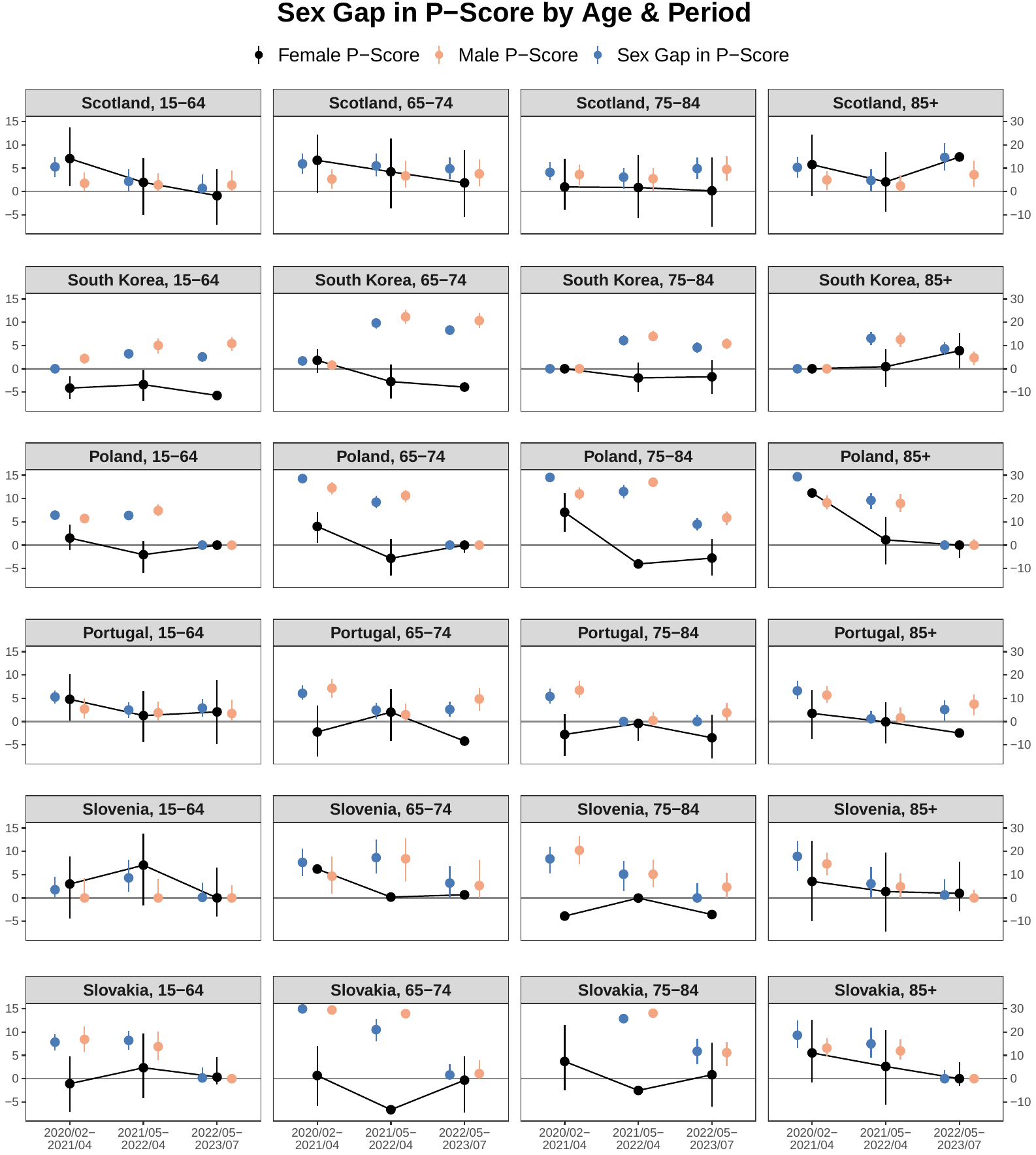

Figure S 21. Excess death P-score by sex (left y-axis) and the sex gap in P-score (male minus female, right y-axis), shown by age group, period, and country [Poland, Portugal, Scotland, Slovakia, Slovenia and South Korea]. Part 5 of 6.

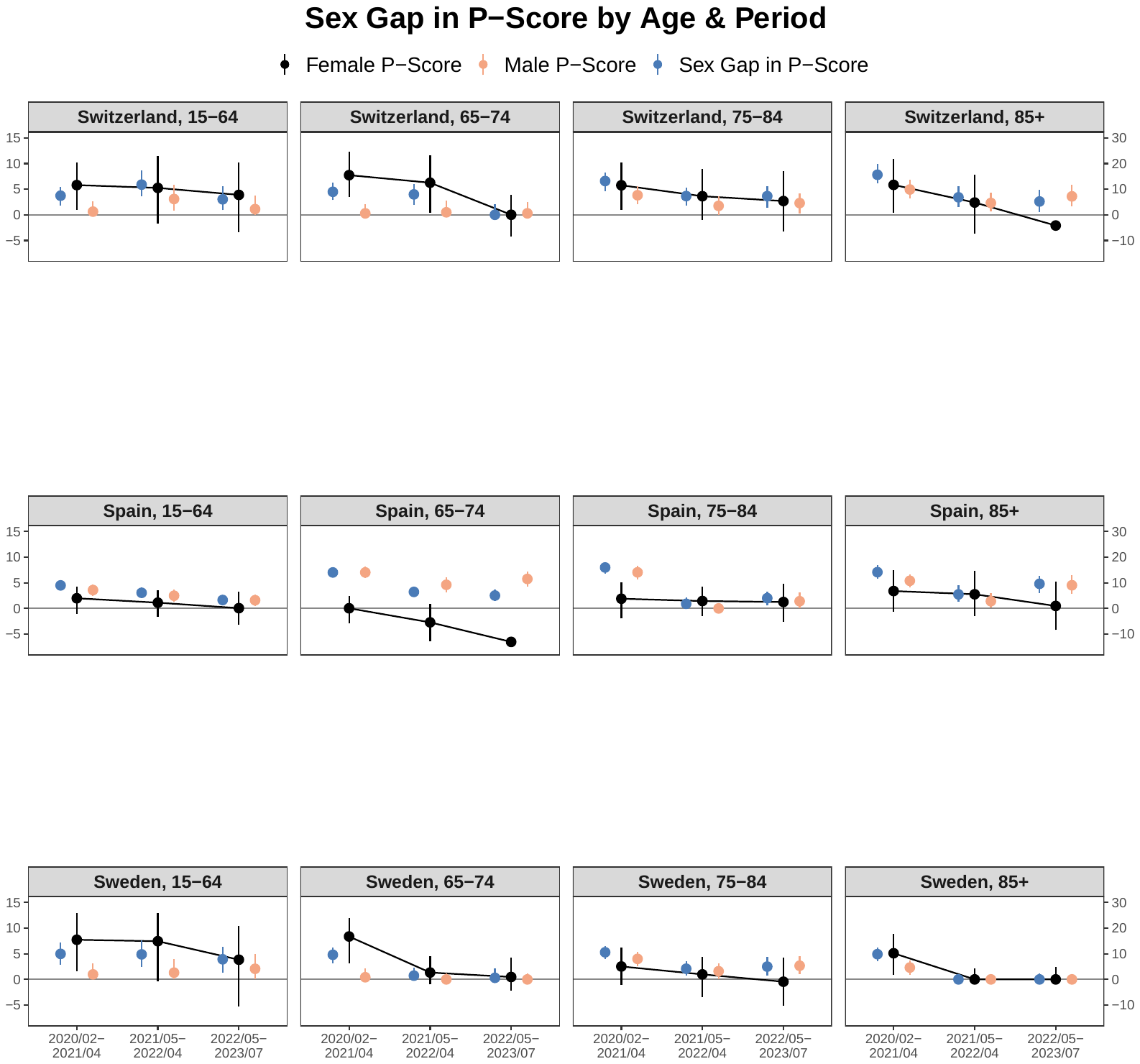

Figure S 22. Excess death P-score by sex (left y-axis) and the sex gap in P-score (male minus female, right y-axis), shown by age group, period, and country [Spain, Sweden, and Switzerland]. Part 6 of 6.

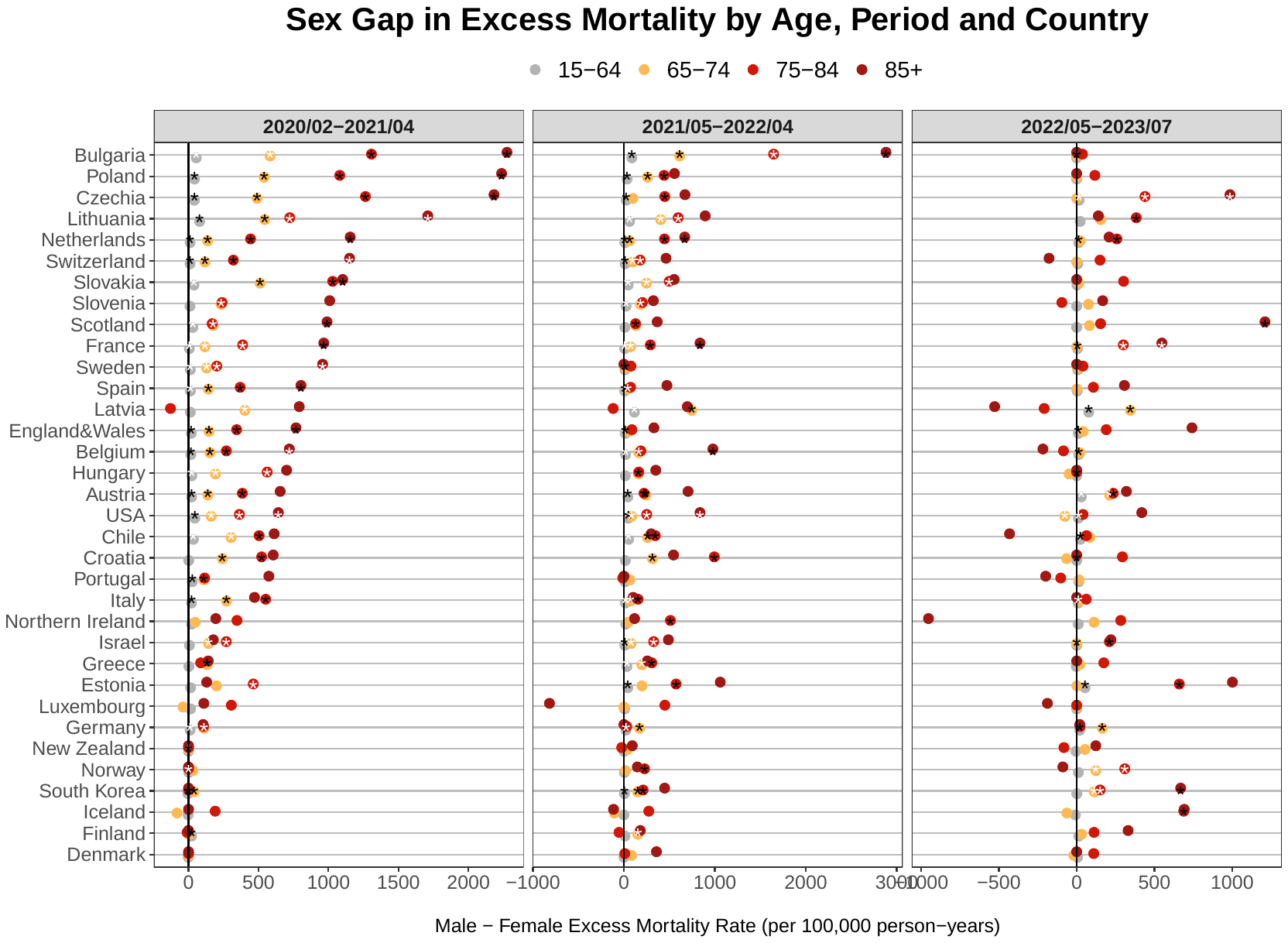

Figure S 23. Sex gap (male-female) in excess all-cause mortality death rate (per 100,000 person-years) by age group, period and country.

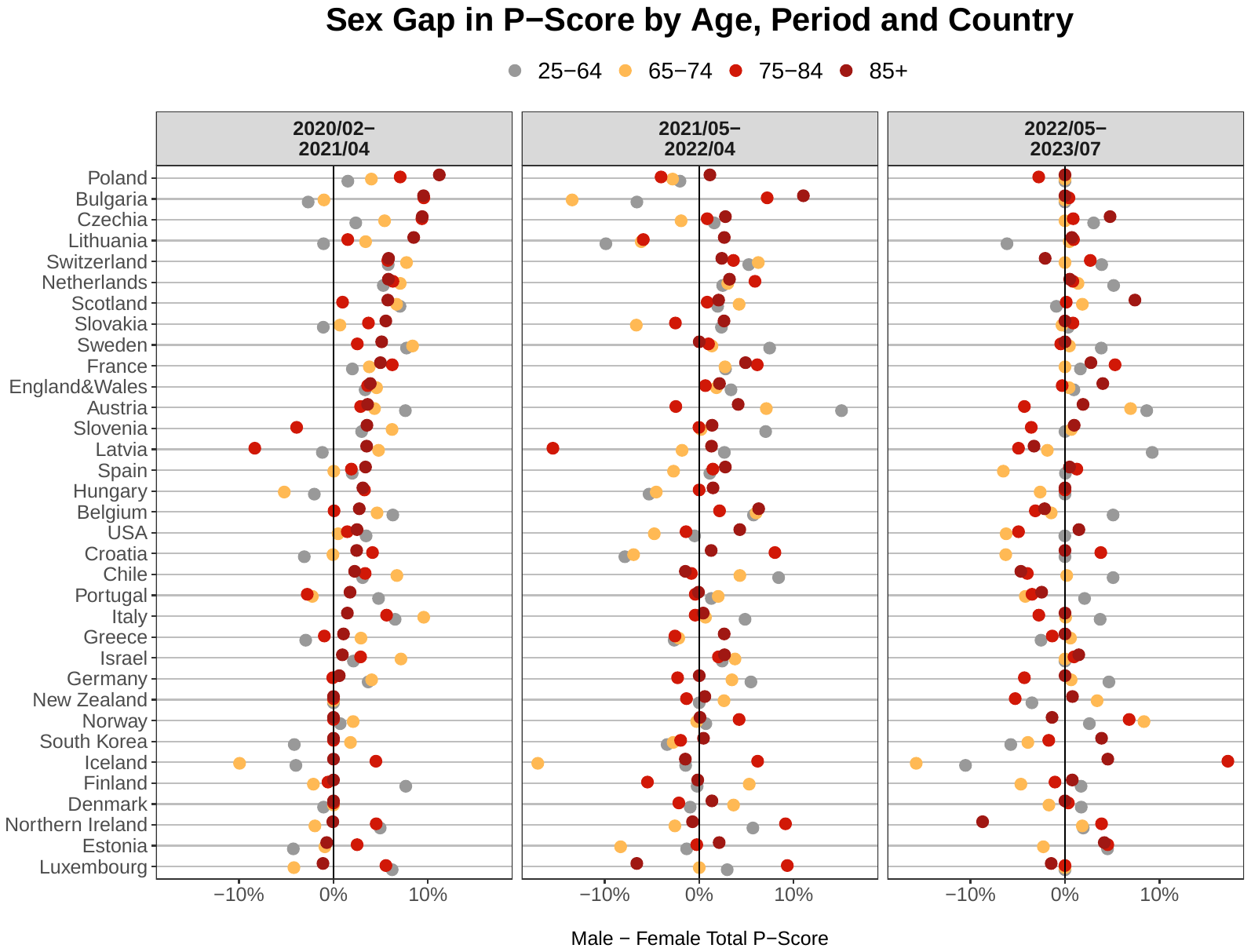

Figure S 24. Sex gap (male-female) in P-score by age group, period and country.
